# Unlocking Esophageal Carcinoma’s Secrets: An integrated Omics Approach Unveils DNA Methylation as a pivotal Early Detection Biomarker with Clinical Implications

**DOI:** 10.1101/2023.09.26.23296198

**Authors:** Akbar Ali, Li Zhang, Hong-Sheng Liu

## Abstract

Esophageal carcinoma (EC) ranks among the top six most prevalent malignancies worldwide with a recent surge in incidence. An innovative integrated omics technique is presented for discerning the two primary types of esophageal carcinoma (EC) AND Squamous cell carcinoma and adenocarcinoma. Utilizing The Cancer Genome Atlas (TCGA) data via Bioconductor, the research integrated DNA methylation and RNA expression analyses for esophageal cancer (ESCA). Key findings revealed DNA methylation’s pivotal role in ESCA progression and its potential as an early detection biomarker. Significant disparities in methylation patterns offered insights into the disease’s pathogenesis. A comparison with the TCGA Pan-Cancer dataset using Bioconductor tools enriched the understanding of ESCA genomics. Specifically, 131,220 hypomethylated probes were detected in tumors compared to 6,248 in healthy tissues. Additionally, 42,060 probe-gene pairs linked methylation variations to expression alterations, with 768 hypomethylated motifs identified. Thirteen of these motifs emerged as potential diagnostic markers. Transcription factor analyses spotlighted crucial regulators, including NFL3, ATF4, JUN, and CEBPG, revealing intricate regulatory networks in ESCA. Survival statistics further correlated clinical factors with patient longevity. This research recommends an innovative approach to identifying oesophageal abnormalities through DNA methylation and gene expression mechanisms. Research suggests DNA methylation may serve as an early detection biomarker, aiding in identifying esophagus cancer prior to more advanced stages.

## 2 Introduction

**Figure 1.1.**
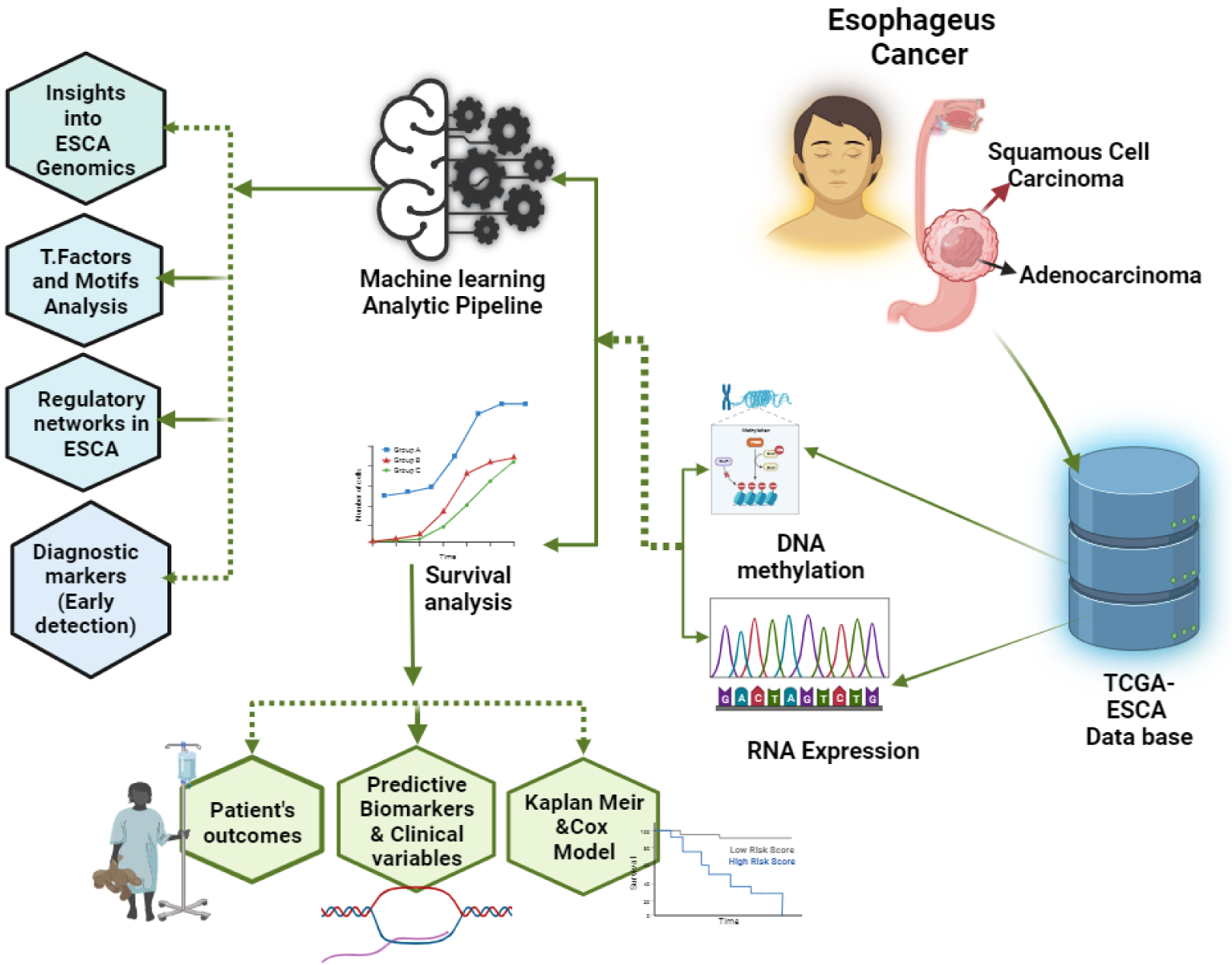
Graphical abstract of Unlocking Esophageal Carcinoma’s Secrets: An Integrated Omics Approach Unveils DNA Methylation as a Pivotal Early Detection Biomarker with Clinical Implications

Esophageal cancer is one of the foremost global causes of mortality [1]. There are two types of ESCA cancer: EAC (esophageal adenocarcinoma (and squamous cell carcinoma (ESCC) [2]. EAC prevails in Western populations, whereas ESCC predominates in Asian nations, particularly China, with an incidence rate of 88.84 [3].

At the core of esophageal cancer is DNA methylation, a pivotal epigenetic modification in mammalian genomes renowned for its manifold applications, including genome identification and gene expression control[3], probing transcriptional enhancers, and other cis-regulatory modules (CRMs). While DNA methylation analyses conventionally focused on gene promoter regions [4]. The exploration of the influence of enhancer regions was initially a less-trodden path. Only through systematic and impartial analyses of DNA methylation in human cells did enhanced regions reveal themselves in a cell-specific context. The advent of target bisulfite sequencing corroborated the cell-type-specific demethylation of enhancers in ENCODE project[5]. Recent investigations into cancer tissues have revealed the potential of DNA methylation profiles to identify cancer-specific enhancers and binding sites for transcription factors. Notably, current studies have indicated that enhancements in DNA methylation within enhancer elements exhibit a significantly stronger predictive power for changes in target gene expression in cancer than those observed within promoters 3. Encouraged by the recognition and discovery of tumor enhancers, transcription factor binding sites (TFBSs), and other cis-regulatory modules (CRMSs) in DNA methylation information and the positive association between DNA methylation and objective gene expression in tumors, the Enhancer by Linking Methylation/Expression relationship package has been developed 4–6. Advanced tools, such as TENET and RegNetDriver, have been employed within this package, along with codes for conducting cancer network analyses[6]. The latest iteration, Enhancer by Linking Methylation/Expression relatiosnhip V.2, introduces advanced statistical features, including integration with the TCGABiolinks package to facilitate cohort selection and data import from the NCI Genome Data Commons, utilization of the MultiAssayExperiment (MAE) gold standard data structure to support both Infinium HMR450 and RNA-seq arrays, integration with the TCGABiolinksGUI tool, enhanced testing and exception handling, and comprehensive presentation of results in an HTML file containing data tables, source code, and figures [7]

The emerging Elmer package presents a remarkable capability to dissect TCGA cancer data to unravel distinctive molecular networks. We harnessed TCGA ESCA cancer data to meticulously probe the methylation and expression datasets within the scope of our project. This potent package finds its home in the Bioconductor R environment establishing a user-friendly avenue for accessing its features. Elmer’s conceptualization centers on the comprehensive exploration of DNA metadata and gene expression data derived from tissues. This tailored framework facilitates the detection of distant probes situated apart from genes and subsequently compares them with neighboring genes. Through this dynamic process, Enhancer by Linking Methylation/Expression relationship, unveils an array of transcriptional targets, illuminating a web of regulatory interactions that might otherwise remain concealed using conventional methods. By embracing the Elmer package, researchers can navigate the complexities of TCGA cancer data with heightened efficiency. This innovative tool catalyzes a streamlined analysis process, aiding in exploring the intricate molecular landscapes that characterize cancer. [8].

Demonstrating survival predictions using TCGA clinical data is pivotal as it sheds light on patient prognosis and outcome metrics [11]. These survival predictions can be contingent on factors such as disease progression, mortality, or even encompassing cases without disease specificity. Identifying potential therapeutic targets and enhancing prognostic accuracy are essential for medical practitioners. Genome-wide assessment of messenger RNA transcripts has emerged as a potent approach for identifying gene expression. This discerning capability positions these genetic markers as indispensable indicators of clinical outcomes, particularly within the domain of survival analysis in cancer research [9]. This significant breakthrough equips researchers and physicians with tools to delve deeper into patient survival dynamics, facilitating informed medical decisions and customizing patient care strategies.[9, 10]

## 3 Material and method

**Figure 3.1.**
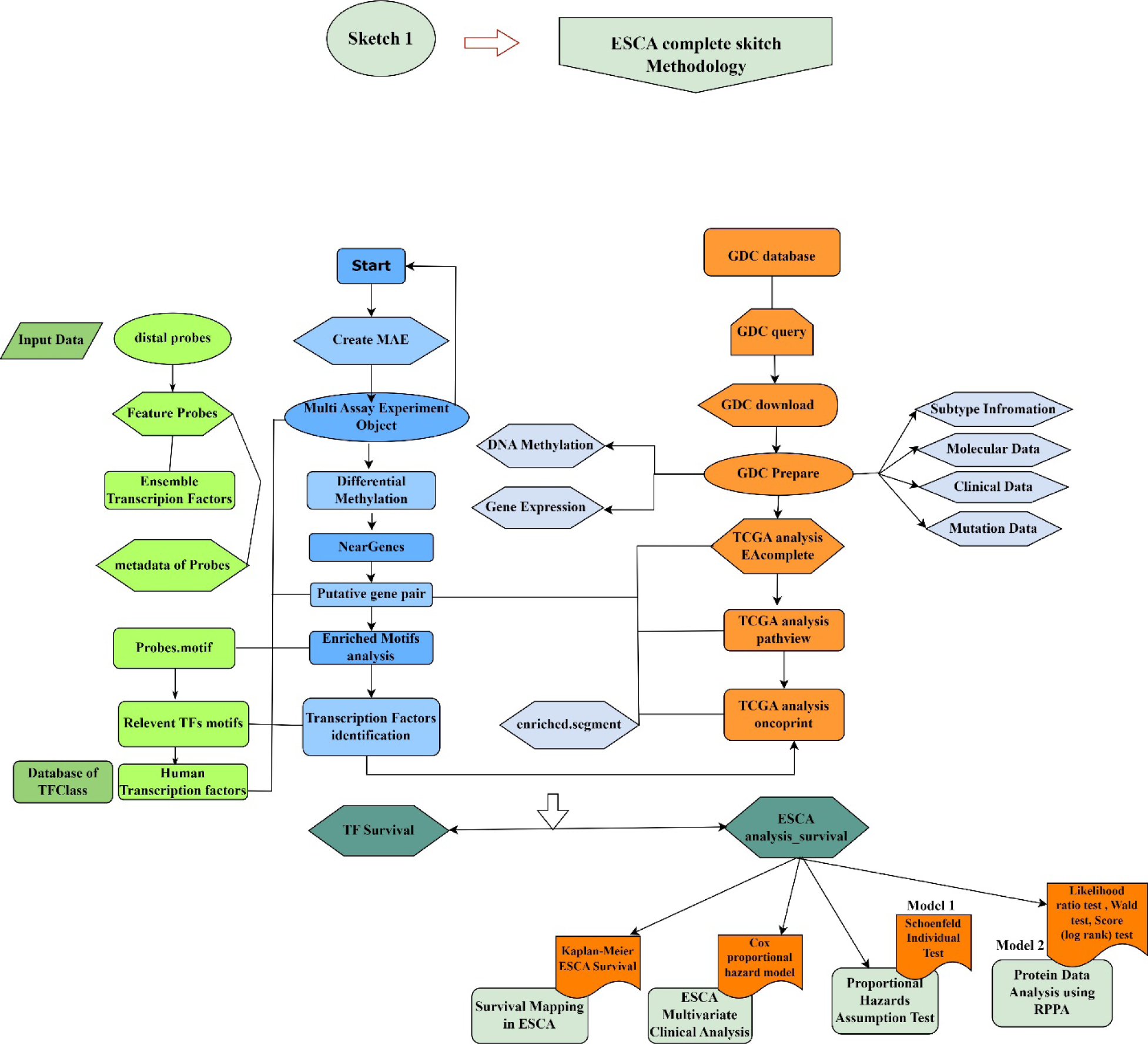
Pathway Diagram of Esophageal Cancer (ESCA)

The flowcharts outlined the methodology from DNA collection to using R-based unsupervised machine learning for identifying methylation patterns in tumors versus normal tissues. They highlighted hypo and hypermethylated probes and key transcription factor motifs within enhancer regions. These charts not only visualized our process but also enhanced the study’s credibility and clarity, illustrating the role of DNA methylation in esophageal cancer.

### 3.1 Data Nexus and R Code Library

The open-source Bioconductor repository at http://bioconductor.org/packages/ELMER/ provides access to source code versions for R, particularly R version 4.3.1 or higher. This package relies on two main dependencies: (I) TCGA, which simplifies the retrieval of cancer data (specifically ESCA) from the NCI’s Genomic Data Common (GDC), and (II) MultiAssayExperiments, which offers a structured and organized data framework within the R environment. It is recommended that a minimum of 16 GB of RAM be allocated for the optimal analysis of cancer data. Notably, no definitive model for DNA methylation and RNA expression analysis exists, but following these suggested specifications can significantly enhance the efficiency and accuracy of data processing[6]. The Enhancer Linking by Methylation/Expression Relationships package is an algorithmic Bioconductor tool that integrates DNA methylation profiles and gene expression patterns to infer multilevel cis-regulatory networks in human tissues[11]. ELMER identifies transcriptional target sites linked to distant probes and local gene expression, thereby unveiling changes in transcriptional enhancers and cis-regulatory modules in tumor cells and primary disease tissues [6]When applied to TCGA cancer datasets as well as DNA and gene expression datasets, Enhancer linking by methylation/expression relationship illuminates intricate regulatory interactions within molecular data.

### 3.2 Data Preprocessing and Implementation

The Enhancer Linking by Methylation/Expression Relationships analysis was carried out primarily in five steps in R using the Bioconductor package [12]. In this study, we aimed to examine DNA methylation-gene connections using distant probes on the HM450K array in a multi-assay experiment. Our focus was on the identification of distinct distant probe clusters with varying levels of DNA methylation [12, 13]. We combined methylation and expression data to identify genes associated with diverse methylation patterns in remote studies. We also identified enrichment motifs for individual probes to reveal the regulatory elements in gene methylation dynamics[13]. Finally, we aimed to uncover a novel transcription factor that governs DNA methylation by overseeing regulatory region expression. Following these steps, we can uncover the intricate relationships between distant probes, DNA methylation changes, gene expression, and regulatory mechanisms.

### 3.3 Data sets

The TCGAbiolinks package includes the “get-TCGA” feature, which provides an updated method for obtaining TCGA data from various samples, including ESCA and other relevant cancer datasets. [14].The Genome Browser feature operates in two modes, with “hg19” retrieving data from the legacy GDC database and “hg38” acquiring data from the latest GDC harmonized data portal[15].

### 3.4 DNA methylation data

The amalgamation of TCGAbiolinks and Enhancer linking by methylation expression realtionship has enabled the scrutiny of ESCA cancer by employing the most recent TCGA datasets. The acquisition of DNA methylation datasets and their conversion into SummarizedExperiment objects was executed by the “getTCGA” function [16]. In addition, gene expression quantification datasets were obtained. This unsupervised machine learning facilitates the mechanization of extracting relevant information from the GDC website and converting it into a matrix format. The research discovered noteworthy M(M+U) values, which represent methylated and unmethylated allele intensities, augmenting our comprehension of ESCA cancer and demonstrating the advantages of combining TCGAbiolinks and Enhancer Methylation/expression relationship [17, 18].

### 3.5 Gene Expression Data

To precisely evaluate the ESCA cancer information obtained from TCGA, we utilized the TCGAbiolinks R package in R for importing the top-notch Expression data (HTSeq-FPKM-UQ) furnished by the Cancer Genome Atlas (TCGA). We meticulously transformed the data into ELMER matrices, ensuring accuracy at every step, and eventually stored the resultant matrix as an RDA file named “RNA.rda” [19].

### 3.6 Integration with Downstream Tools and statistical Modeling inference

The Bioconductor team created the Summarized Experiment class to aid in the analysis of multiple samples; however, it was limited in its ability to store data from multiple experiments with shared samples. To overcome this, the MultiAssayExperiments class was developed by the MultiAssayTrial group to cater to the pre-processing requirements for integrated genomic analysis tests. [16]. Consequently, the create-MAE service generated a data configuration for Unsupervised machine learning, linking methylation/expression analysis. So this analysis, a MultiAssay experiment containing a DNA methylation matrix or a summary experiment object from the HM450K or EPIC platform was utilized. When TCGAbiolink imported TCGA and other GDC data, an automated data structure was created to empower unsupervised machine learning to handle diverse datasets, including metadata for DNA methylation probes. These metadata comprised aggregated genomic coordinates based on the sample genome and gene annotations sourced from the ENSEMBL database [^8^].

### 3.7 Choosing distal probes

The utilization of default filtering, as presented in, led to the removal of specific probes from the HumanMethylation (EPIC) and Infinium HumanMethylation450 (HM450 array) platforms [20]. default filtering manifests itself. Samples that contained both SNPs near the 3’ ends were classified as masked and, as a result, were excluded from the analysis. Elmer provides access to comprehensive probe metadata. Specifically, when analyzing the distal component, probes located within the +2 kb region surrounding the transcription start sites (TSSs) were disregarded [21].

### 3.8 Unleashing Insights via Unsupervised Data Analysis

This research delves into ESCA cancer data analysis, emphasizing leveraging unsupervised techniques to reveal the underlying patterns. This study initiated a meticulous phase of hard coding, facilitating the distinction between cancerous and non-cancerous samples. Subsequently, the focus transitions to unsupervised analysis aimed at uncovering nuanced variations within specific tumor subgroups [10](). Notably, integrating ELMER version 2 is a pivotal component, presenting a versatile framework applicable to paired datasets [10]. This framework facilitates comprehensive comparisons, encompassing disease versus health scenarios across diverse conditions and differences between untreated and treated specimens. In essence, this investigation stands as a significant stride in ESCA cancer research, employing a synergy of techniques to decode intricate data patterns and potentially catalyze further scientific inquiry.

### 3.9 Proteins’ Impact on Survival: An Extensive Data Analysis Exploration

The ESCA cancer datasets were obtained from the MD Anderson Cancer Center’s TCPA database[20]. Level four of the data analysis involved sourcing pertinent clinical and survival data for the ESCA cancer dataset [22]. This included essential information such as overall survival, disease-specific survival, disease-free intervals, and progression periods, which were then linked to primary RPPA tumor identifiers specific to ESCA cancer 21. To further explore the ESCA survival data, R version 4.3.1. Interactive graphics were created using ggplot2 (v3.2.1) and shiny (v1.4.0) 23. Kaplan–Meier survival curves were used to visualize survival trends [23]. These were generated by selecting the relevant ESCA subtypes, survival rates, and specific protein(s) of interest. This comprehensive analytical approach provides valuable insights into the survival dynamics of patients with ESCA [24]. Our study focused on critical proteins analyzed via immunohistochemistry, which is vital in breast cancer research [25] Specifically, we focus on the ESCA study. We employed Kaplan-Meier analysis to analyze the survival data and utilized R’s survival (v3.1-7) and survminer (v0.4.6) packages. Multiple protein survival analysis was aligned with average protein probe standards, which is similar to gene expression methods [14, 26].

To compute hazard ratios for two-group comparisons, we used Cox proportional hazard regression from the survival package [27]. We used median or optimal cutoffs and Survminer’s optimal cutoff function, based on the log-rank test and tailored for 15% variance, to identify significant biomarkers[28].

## 4 RESULTS

### 4.1 Decoding Epigenetic Changes: Recognizing Differentially Methylated CpGs (DMCs)

The initial stage utilized the (get.diff.meth) function to identify differentially methylated CpGs. In the Unsupervised Machine Learning approach, samples were segregated into groups 1 and 2 based on the DNA methylation beta values for a particular probe. The lowest 20 percent of samples with minimal methylation were handpicked from each group to compare the hypomethylation of Group 1 with that of Group 2. In Unsupervised Machine Learning mode, each selected probe was centered on a distinctive subset of samples, representing an array of molecular subtypes. The unsupervised method functioned with sample sets encompassing 20% [29]. This selection of 20 percent ensured a sufficient number of samples for precise identification of a specific molecular subtype while generating t-test p-values significant for multiple hypothesis corrections[30].

Hypomethylated differentially methylated CpG sites (DMCs) were identified, and a one-tailed t-test was employed to test the Null Hypothesis (μgroup1 μgroup2). The raw p-value represents a fixed number for multiple hypothesis testing procedures that demonstrate a significant p-value and adhere to the Benjamini-Hochberg approach with a default p-value of 0.01[31]. Moreover, only probes with a methylation difference Δ = μgroup1 − μgroup2 greater than 0.03 were chosen 3,28. The approach used to discover hypermethylated DMCs, apart from the different tails in the t-test, was selected and employed in the upper quintile.

**Figure 4.1.**
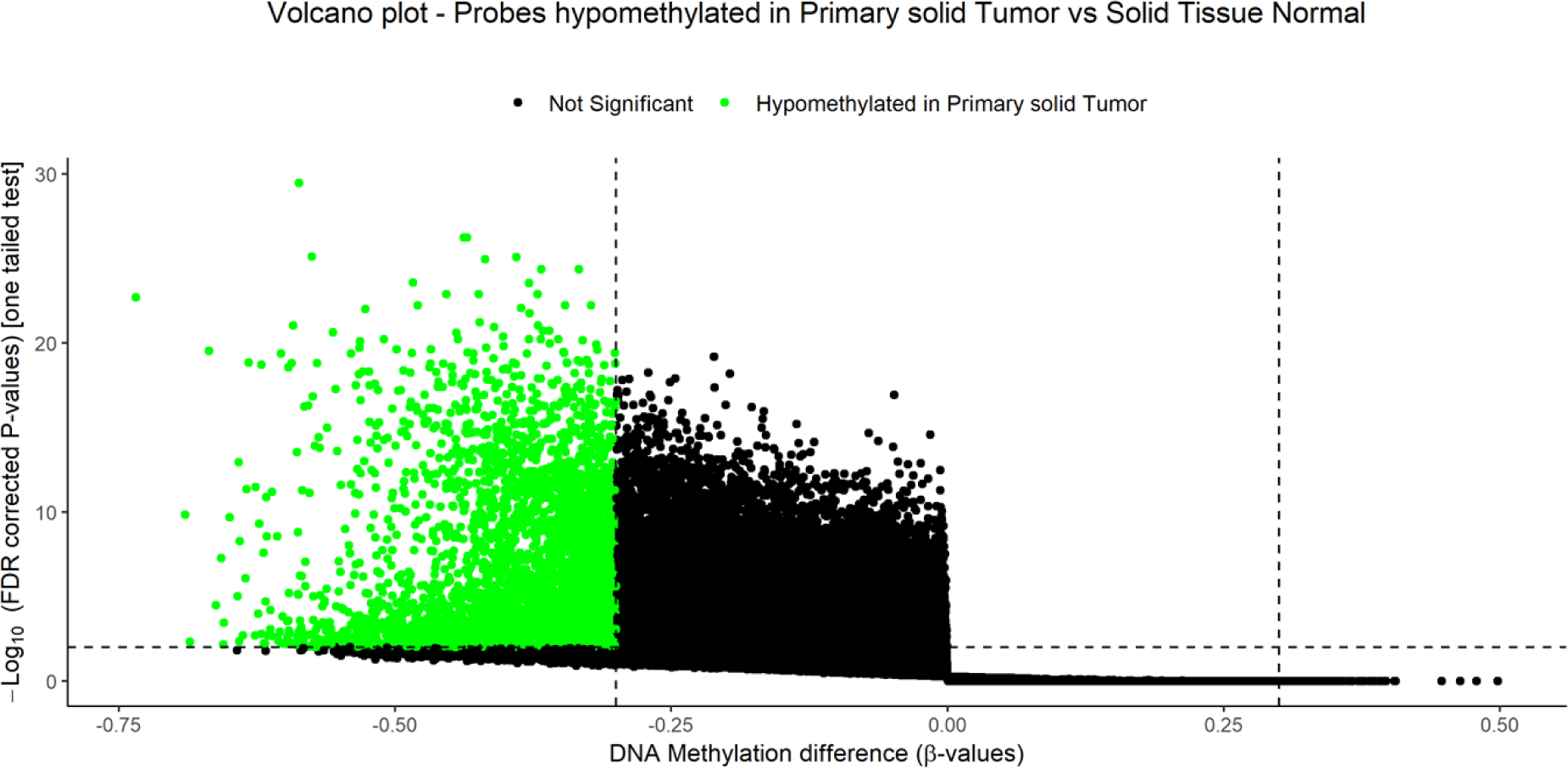
Probes hypomethylation in primary solid tumors in comparison to normal solid tissue

The presented visual representation depicts a volcano plot that effectively highlights hypomethylation probes in primary solid tumors when compared to normal solid tissue. The plot features a scatter plot with an x-axis labeled as “DNA Methylation Difference (β-values)” and a y-axis labeled as “-Log10 (FDR Corrected P-values) [One-Tailed Test].” The plot’s data points are color-coded, with green points indicating significant hypomethylation in primary solid tumors and black points representing non-significant probes. Moreover, the plot includes reference points through the inclusion of dashed vertical (x = 0) and horizontal (y = 1.5) lines.

**Table 4.1.** Data table containing crucial information regarding the probe.

| Probe | p-value | Primary.<br>Solid.Tumor_Minus<br>Solid.Tissue. Normal | adjust.p |
| --- | --- | --- | --- |
| cg00002749 | 1.53E-25 | -0.592 | 9.15E-22 |
| cg00014484 | 1.08E-08 | -0.407 | 2.81E-07 |
| cg00016481 | 6.98E-13 | -0.385 | 5.90E-11 |
| cg00018696 | 1.49E-05 | -0.302 | 1.47E-04 |
| cg00019997 | 1.03E-12 | -0.330 | 8.24E-11 |
| cg00022145 | 0.00114 | -0.387 | 0.0053 |
| cg00026776 | 3.69E-10 | -0.340 | 1.45E-08 |
| cg00028598 | 0.00120 | -0.503 | 0.0055 |
| cg00030476 | 5.86E-04 | -0.319 | 0.0031 |
| cg00033490 | 1.21E-04 | -0.392 | 8.66E-04 |
| cg00035220 | 1.26E-12 | -0.373 | 9.88E-11 |
| cg00039129 | 7.77E-04 | -0.333 | 0.0039 |
| cg00050692 | 2.31E-07 | -0.464 | 4.03E-06 |
| cg00066854 | 4.67E-04 | -0.421 | 0.00263 |
| cg00069815 | 9.48E-17 | -0.552 | 2.55E-14 |
| cg00085256 | 3.16E-13 | -0.305 | 2.94E-11 |
| cg00088507 | 2.40E-15 | -0.367 | 4.27E-13 |
| cg00091146 | 1.09E-04 | -0.385 | 7.96E-04 |
| cg00107916 | 1.28E-04 | -0.406 | 9.09E-04 |
| cg00108617 | 7.87E-07 | -0.439 | 1.16E-05 |
| cg00108938 | 1.32E-18 | -0.365 | 6.35E-16 |
| cg00128695 | 0.00136 | -0.312 | 0.006 |
| cg00134787 | 0.00103 | -0.434 | 0.004 |
| cg00134934 | 2.02E-13 | -0.304 | 1.97E-11 |
| cg00136968 | 2.05E-05 | -0.315 | 1.93E-04 |
| cg00138710 | 2.30E-17 | -0.347 | 7.24E-15 |
| cg00145093 | 1.38E-05 | -0.387 | 1.38E-04 |
| cg00145961 | 3.79E-07 | -0.417 | 6.17E-06 |
| cg00146565 | 4.81E-24 | -0.401 | 1.62E-20 |
| cg00180684 | 3.82E-16 | -0.393 | 8.60E-14 |
| cg00186580 | 0.002557 | -0.368 | 0.009 |
| cg00190835 | 3.50E-21 | -0.335 | 3.76E-18 |
| cg00192819 | 0.00121 | -0.414 | 0.0055 |
| cg00205332 | 6.99E-18 | -0.350 | 2.64E-15 |
| cg00205786 | 0.00191 | -0.300 | 0.0078 |

Table 4.1: The data table comprises crucial information regarding the probe p-value, p-value adjustment, beta value, and primary. Notably, the rows in the table featuring a p-value below 0.05 hold significance, as they provide robust evidence against the null hypothesis and point towards the truth of the alternative hypothesis. Among the top 19 probes, cg00002749 and cg00020003 had the highest adjusted p values. In contrast, probes cg00016481, cg00020001, and cg20007 yielded high p-values.

### 4.2 Uncovering Potential Probe-Gene Pairs: A Quest for Identification

The initial step involved employing the get function to link target genes with altered expression to distal probes with altered methylation. Default target genes were established for the samples in question. It is possible to regulate the reverse correlation between methylation of a specific distal probe and the expression of the 10 neighboring upstream and downstream genes. This approach is similar to the primary strategy utilized in ELMER version 1; however, we are presently systematically utilizing the Biomart package to import all gene annotations. [32]. This methodology affords the ability to employ any preferred annotations, with the default Ensemble annotations employed. The M group lacking supervision exhibited the highest methylation of 20%, while the U group exhibited the lowest 20%. We chose the selected genes and the distance between the probe gene, which is defined as the distance from the transcription start site indicated by the ENSEMBL gene-level annotation through the R version/Bioconductor package [33]. Statistical tests were subsequently applied to each probe. For the probe-gene pairs, we utilized the Mann-Whitney U test, with the null hypothesis of total gene expression being equal to group U. As such, we selected solely those x random samples from an interpreted transcription start site that was distal to 2 kb to draw the null hypothesis probes from a similar set as the sample tested [6]. Therefore, we tested the unsupervised model for each pair of probe genes. The raw p-value (Pr) was balanced by the Benjamin-Hochberg method for the hypothesis. We generated a schematic diagram for a probe containing the next 20 adjacent genes, and the gene significantly bound to the probe was labeled in red. Additionally, we produced a table to indicate information about probes.

**Figure 4.2.**
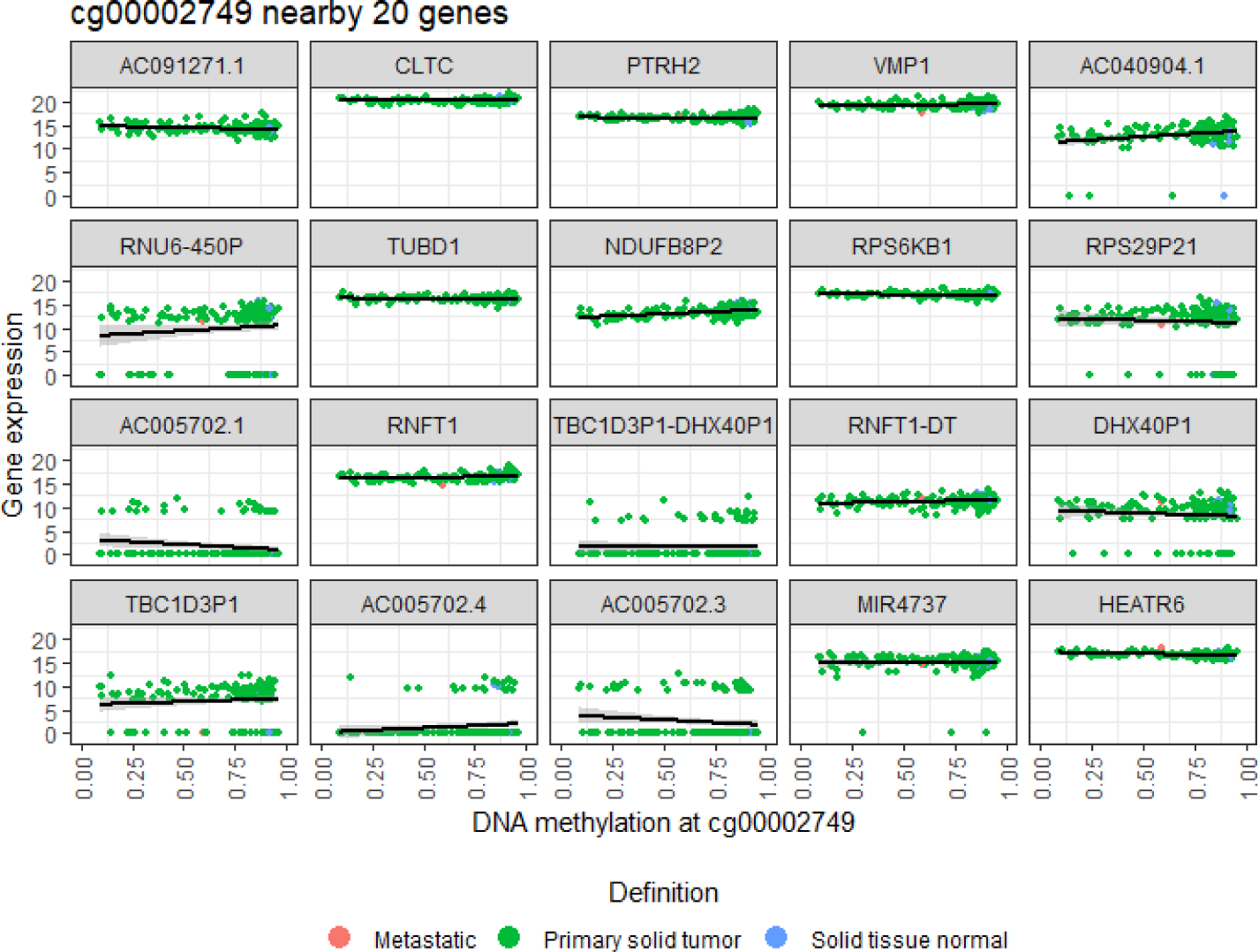
Scatter diagrams were employed to establish a correlation between the methylation of the cg00002749 and Gene expression.

**Figure 4.3.**
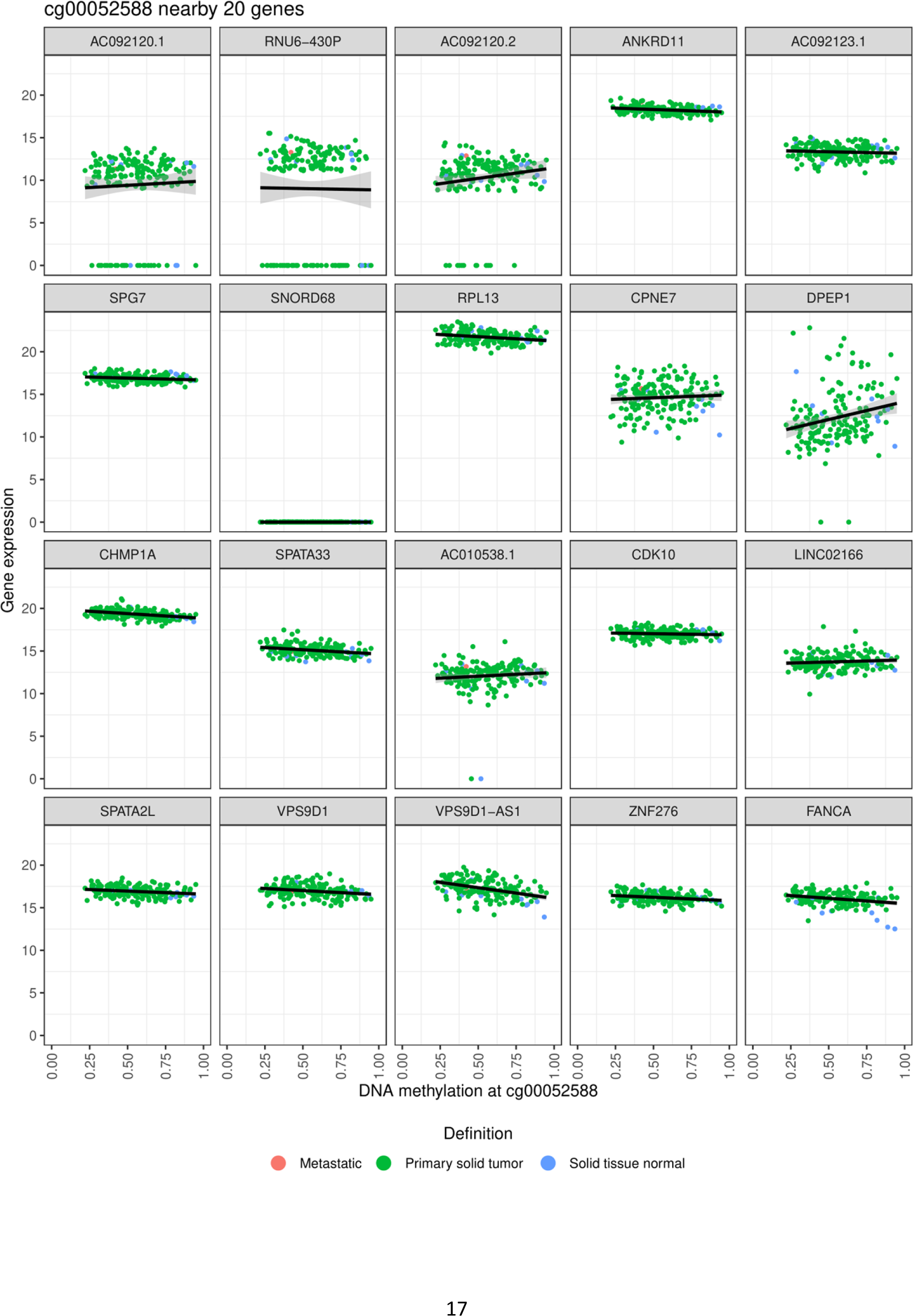
Scatter diagrams were employed to establish a correlation between the methylation of the cg00005258 an d Gene expression.

**Figure 4.4.**
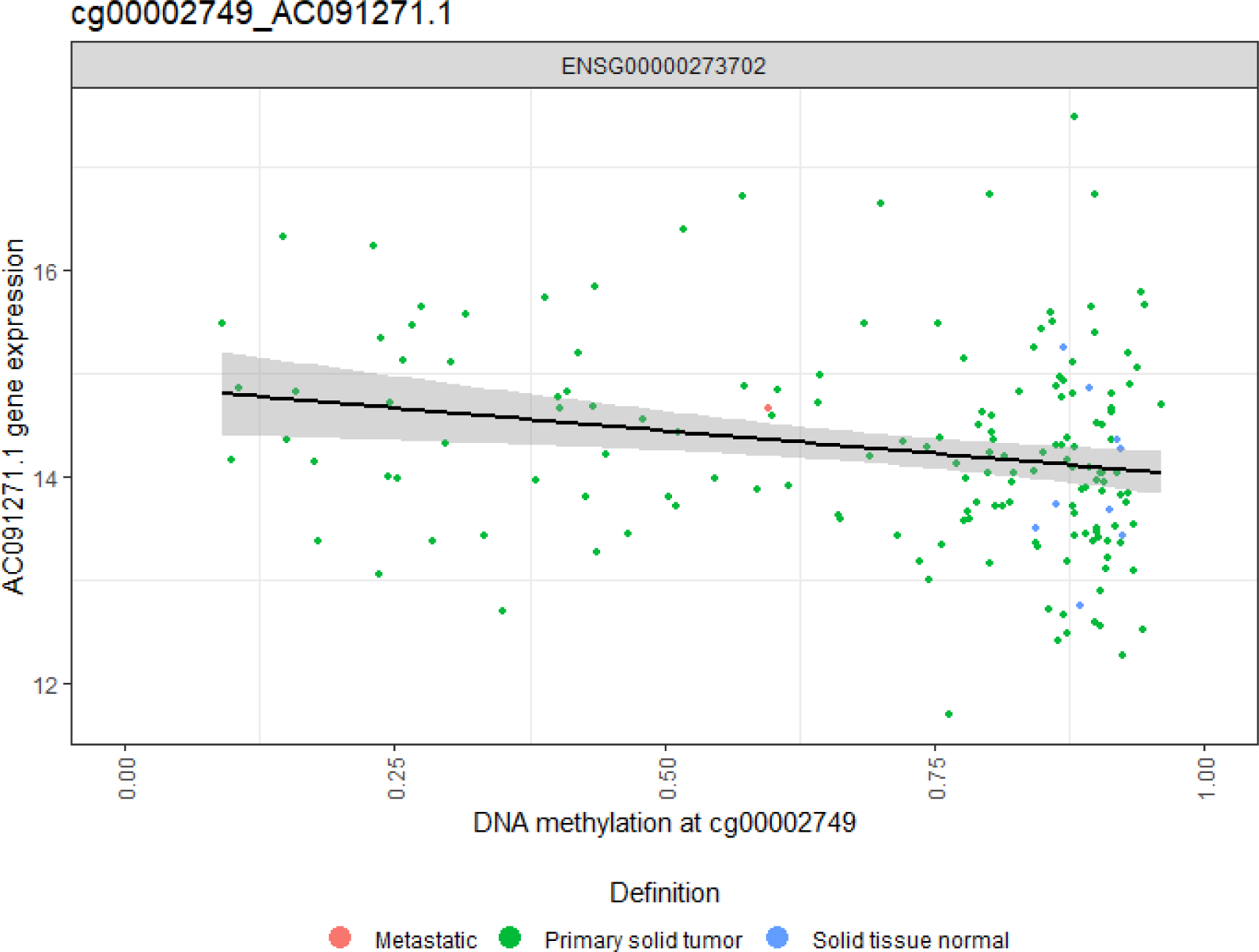
scatter plots were employed to illustrate the methylation of the different probes in ESCA samples.

**Figure 4.5.**
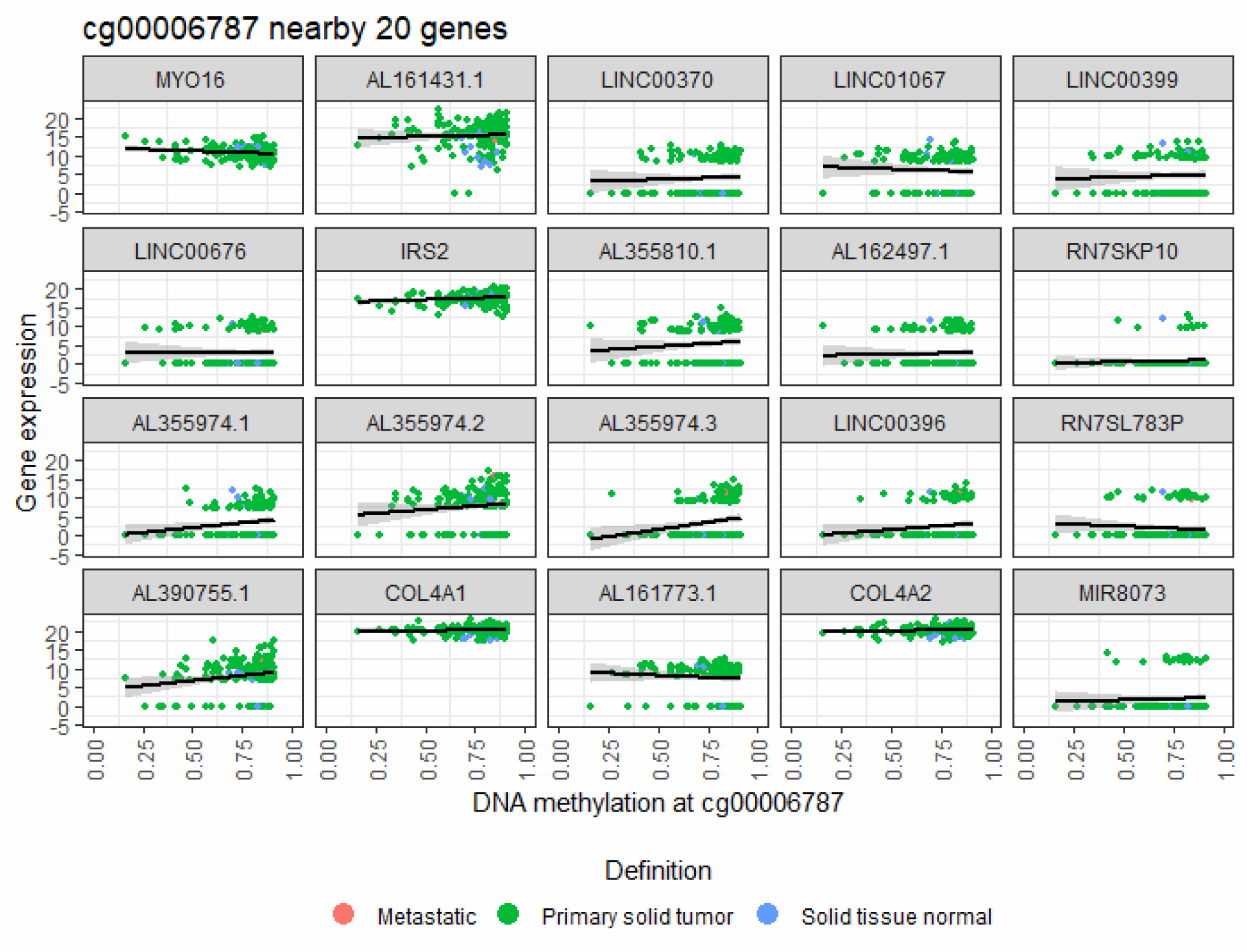
Scatter diagrams were employed to establish a correlation between the methylation of the cg0000687 and Gene expression.

**Figure 4.6.**
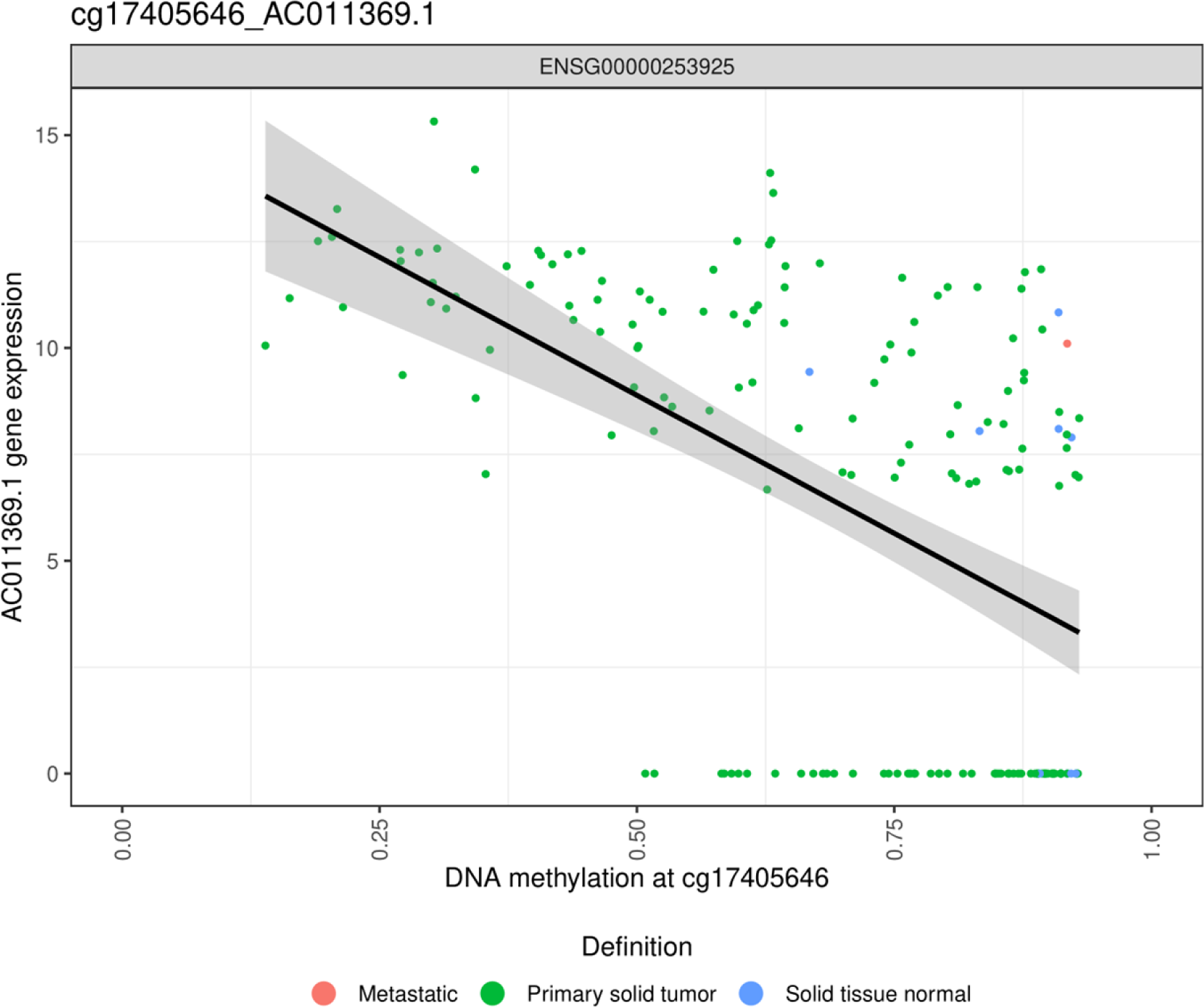
scatter plots were employed to illustrate the methylation of the different probes in ESCA samples.

The study utilized scatter diagrams to elucidate the correlation between the methylation of the cg00002749 probe in ESCA samples and the expression of the adjacent gene. In parallel, the diagrams showcased the methylation of the cg00002749 in ESA samples juxtaposed with the A091271.1 gene expression. Notably, the visualization adeptly portrayed the expression patterns of twenty genes, including CLTC, PTHR2, VMP1, RPS6KB1, RNF1, TBC1D3P1-DH, RNF1- DT, DHX40P1, MIR4737, HEATR6, and more across diverse tissue types. Each sub-chart dedicated to a gene mapped the x-axis to DNA methylation and the y-axis to gene expression. This representation shed light on the interplay between DNA methylation and gene expression in various tissues, offering valuable perspectives into ailments like cancer and paving the way for precision medicine. Moreover, the plots were enriched with lines labeled with gene IDs such as “ACO2102.1”, “RNU6-430P”, and “ANKRD11”. With axes labeled “log2 fold change” and “expression level”, the diagram efficiently juxtaposed gene expression across distinct conditions.

**Figure 4.7.**
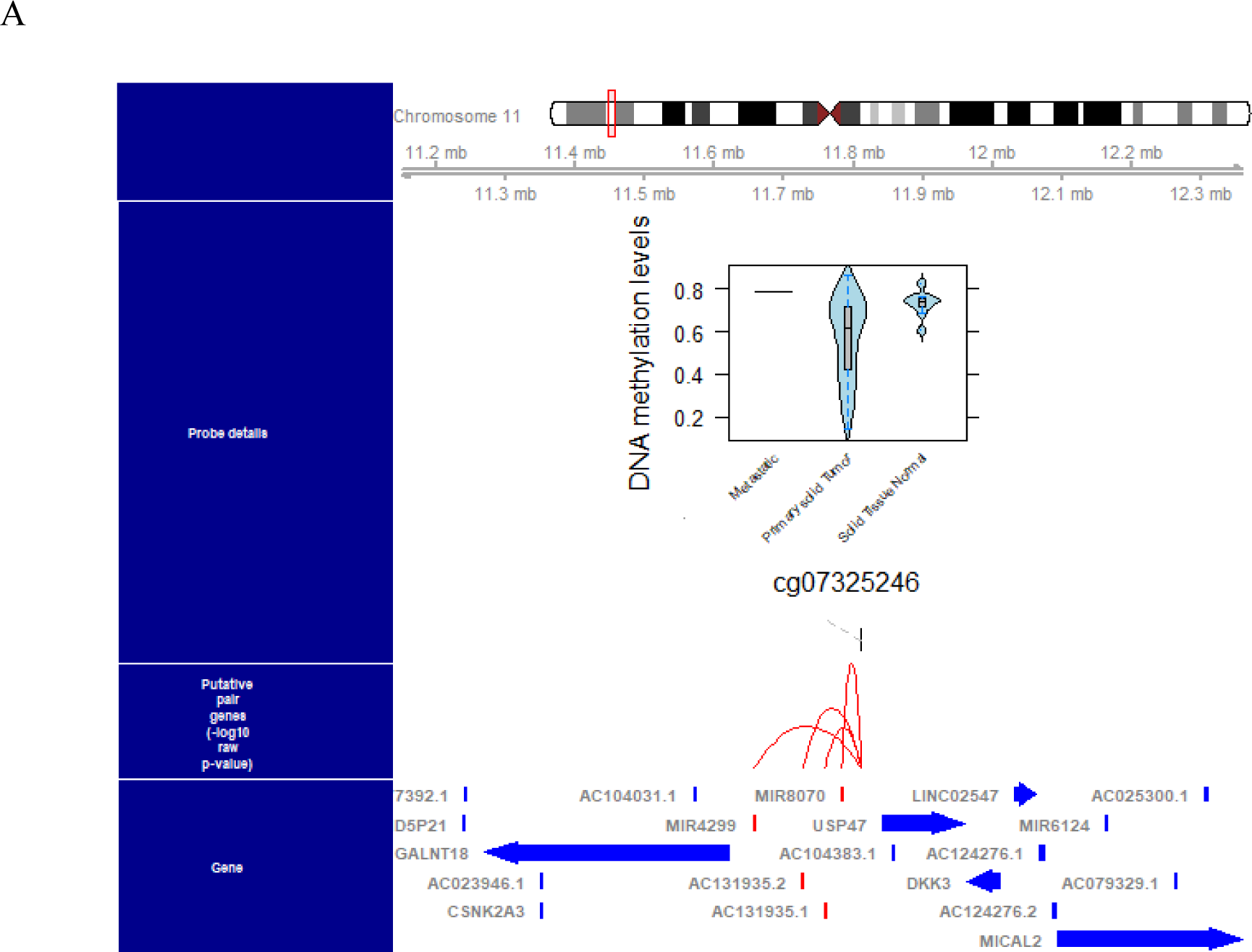
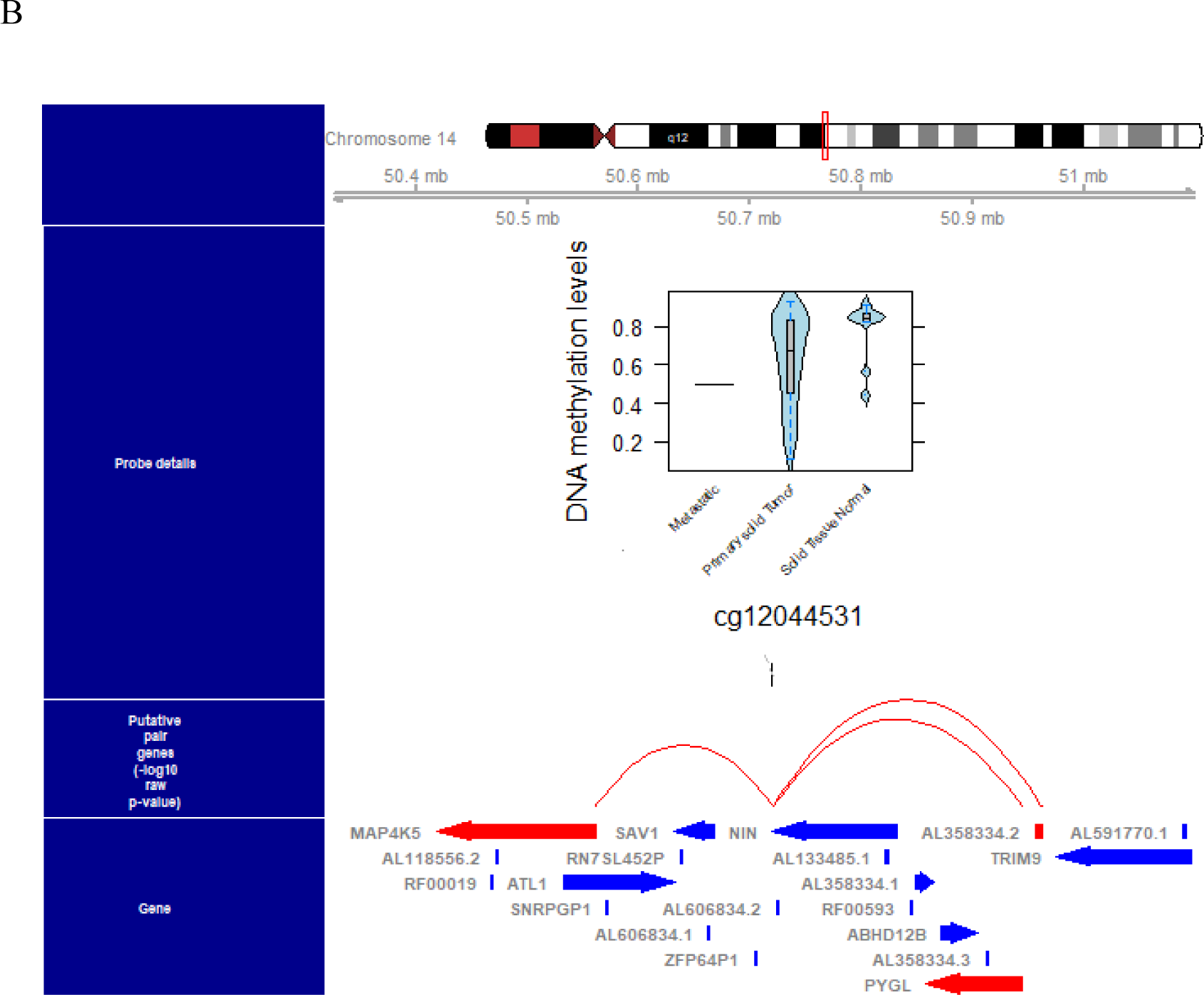
A schematic plot was generated by A and B for a single probe, cg07325246, which included approximately 20 adjacent genes.

#### 3.2.1 Proximal Genes

A schematic plot was created for probe cg07325246, showing nearby genes. The plot highlights the gene linked to the probe in red and the probe in blue. It offers a comprehensive view of DNA microarray analysis, with a chromosome map, DNA methylation levels, and a gene information table. A red line on the map marks the probe data’s location. The table lists gene names with corresponding probe data. The image is labeled “cg07325246”.

### 3.3 Performing motif enrichment analysis on the specific probes and identifying master regulator transcription factors (TFs)

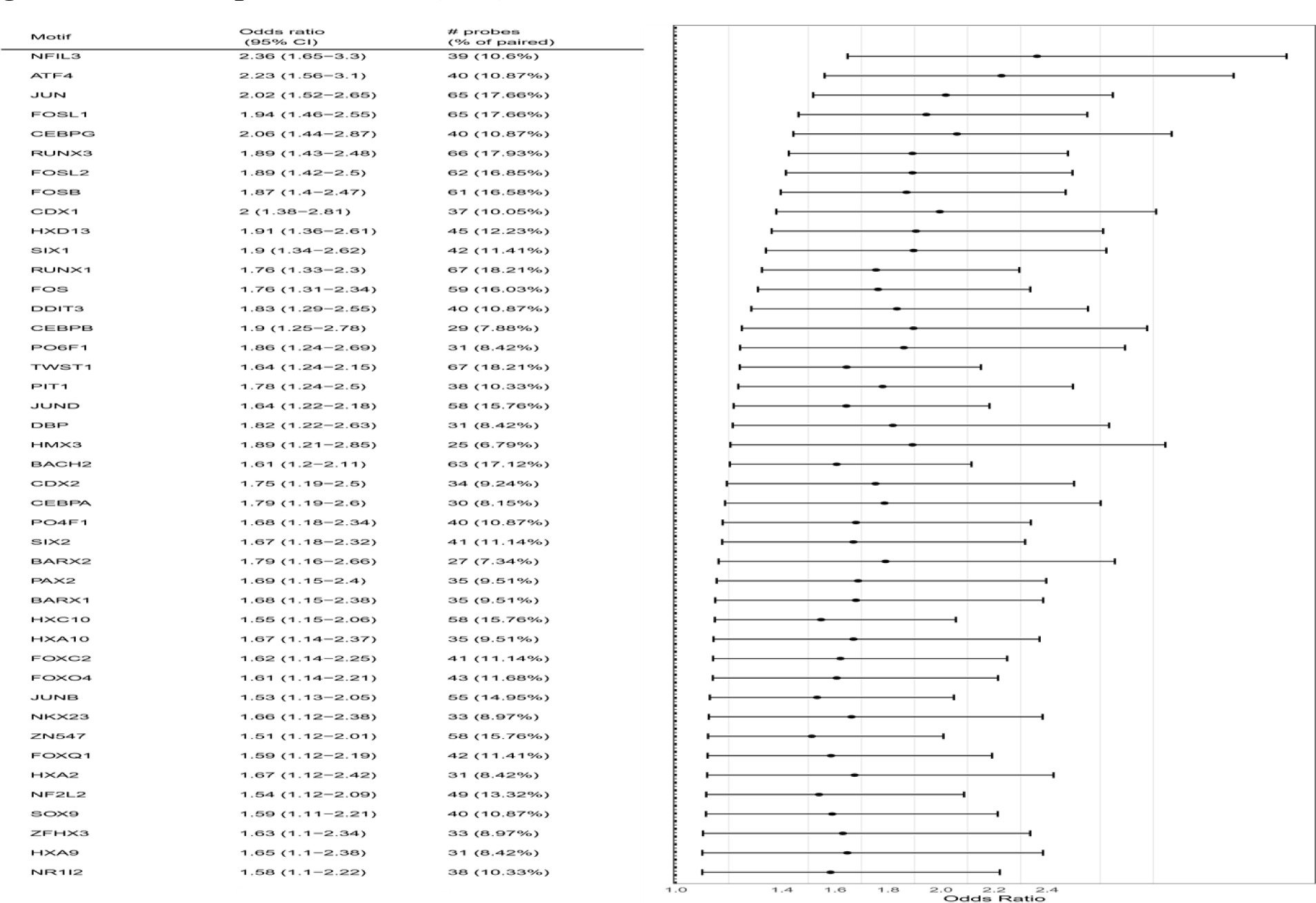

A motif enrichment analysis was conducted on probe-gene pairs using HOMER (Hypergeometric Optimization of Motif EnRichment) with HOCOMOCO v11. This analysis identified enriched motifs and potential upstream transcription factors (TFs) within ±250bp around each probe. The Fisher test was used to determine the enrichment level, and multiple test correction was performed using the Benjamini-Hochberg method. Sets of probes were considered enriched if the 95% confidence interval of the probability ratio was less than 1.1 and if the number of enriched probes was more significant than ten or the false discovery rate was less than 0.05.34

**Figure 4.8.**
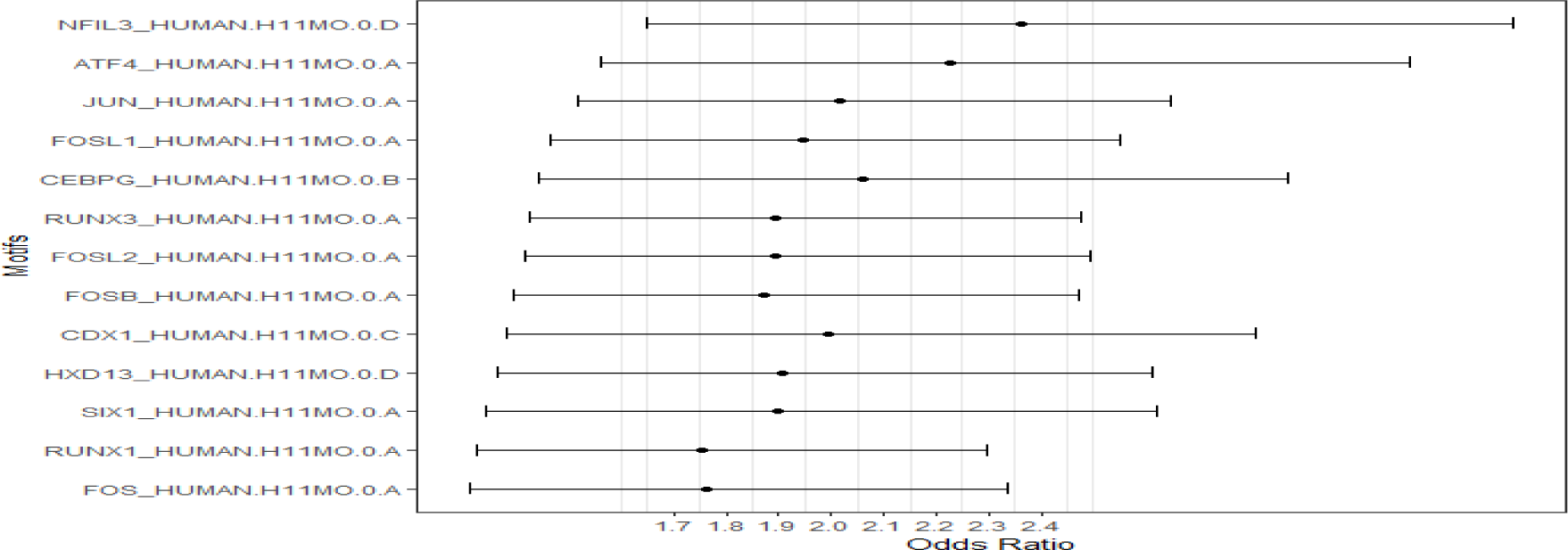
A and B depiction of the predominant motifs utilized for unsupervised analysis of TCGA ESCA cancer.

A figure 4.8 displaying the motifs used for unsupervised analysis of TCGA ESCA cancer data sets is shown. The figures present ratios of probabilities for specific attributes that meet certain criteria. The scales and numbers indicate the corresponding confidence intervals for odds ratio. Motifs with a confidence interval lower boundary exceeding the number by more than 1 are considered enriched. The most significant motifs, including NFL3, ATF4, JUN, CEBPG, and CDX1, have been identified via unsupervised mode analysis of ESCA.

**Table 4.2.** Indicates the top 24 hypo methylated enrichment motifs, their statistical values, and the Transcription family.

Table 4.2 showcases motif enrichment analysis results, revealing DNA sequence motifs’ links to gene regulation. Rows contain details on enriched gene probes, odds ratio range, transcription factor family, and p-values. Notable motifs like NFIL3_HUMAN H11MO.0.D, ATF4_HUMAN H11MO.0.A, and JUN_HUMAN H11MO.0.A, among others, suggest strong gene regulation connections and potential master regulator roles. These insights shed light on gene regulatory mechanisms and disease associations and offer avenues for further exploration and therapy.

**Table 4.3.** Table indicates the 20 hypo methylated enrichment motifs along with their statistical values and Transcription family.

| **Motif** | **No**<br>**of**<br>**Probes** | **% Of**<br>**Probes** | **lowerOR** | **upperOR** | **OR** | **p.value** | **FDR** | **TF family** |
| --- | --- | --- | --- | --- | --- | --- | --- | --- |
| **NFIL3** | 39 | 0.105 | 1.647 | 3.301 | 2.361 | 4.1826E-06 | 0.00026 | C/EBP |
| **ATF4** | 40 | 0.108 | 1.561 | 3.102 | 2.227 | 1.5442E-05 | 0.00066 | ATF-4 |
| **JUN** | 65 | 0.176 | 1.518 | 2.647 | 2.017 | 2.0007E-06 | 0.00022 | Jun-related factors |
| **FOSL1** | 65 | 0.176 | 1.463 | 2.551 | 1.944 | 6.804E-06 | 0.00034 | Fos-related factors |
| **CEBPG** | 40 | 0.108 | 1.443 | 2.868 | 2.059 | 8.7204E-05 | 0.00224 | C/EBP-related |
| **RUNX3** | 66 | 0.179 | 1.426 | 2.478 | 1.892 | 1.0577E-05 | 0.00050 | Runt-related factors |
| **FOSL2** | 62 | 0.168 | 1.415 | 2.495 | 1.892 | 1.877E-05 | 0.00075 | Fos-related factors |
| **FOSB** | 61 | 0.165 | 1.395 | 2.469 | 1.869 | 2.7424E-05 | 0.00096 | Fos-related factor |
| **CDX1** | 37 | 0.100 | 1.379 | 2.810 | 1.995 | 0.00025159 | 0.00436 | HOX-related factor |
| **HXD13** | 45 | 0.122 | 1.361 | 2.610 | 1.905 | 0.00016517 | 0.00363 | HOX-related factor |
| **SIX1** | 42 | 0.114 | 1.340 | 2.622 | 1.896 | 0.00025462 | 0.00436 | HD-SINE factors |
| **RUNX1** | 67 | 0.182 | 1.325 | 2.295 | 1.755 | 9.2012E-05 | 0.00226 | Runt-related factors |
| **FOS** | 59 | 0.160 | 1.309 | 2.336 | 1.762 | 0.00019854 | 0.00405 | Fos-related factors |
| **DDIT3** | 40 | 0.108 | 1.285 | 2.553 | 1.833 | 0.00073152 | 0.01038 | C/EBP-related |
| **CEBPB** | 29 | 0.078 | 1.249 | 2.776 | 1.896 | 0.00192018 | 0.02001 | C/EBP-related |
| **PO6F1** | 31 | 0.084 | 1.242 | 2.693 | 1.860 | 0.00192114 | 0.02001 | POU domain factor |
| **TWST1** | 67 | 0.182 | 1.241 | 2.149 | 1.644 | 0.00049607 | 0.00796 | Tal-related factors |
| **PIT1** | 38 | 0.103 | 1.236 | 2.496 | 1.779 | 0.00146004 | 0.01705 | POU domain factors |
| **JUND** | 58 | 0.157 | 1.218 | 2.182 | 1.643 | 0.0009959 | 0.01301 | Jun-related factors |
| **DBP** | 31 | 0.084 | 1.215 | 2.633 | 1.818 | 0.00309502 | 0.02807 | C/EBP |

**Figure 4.9.**
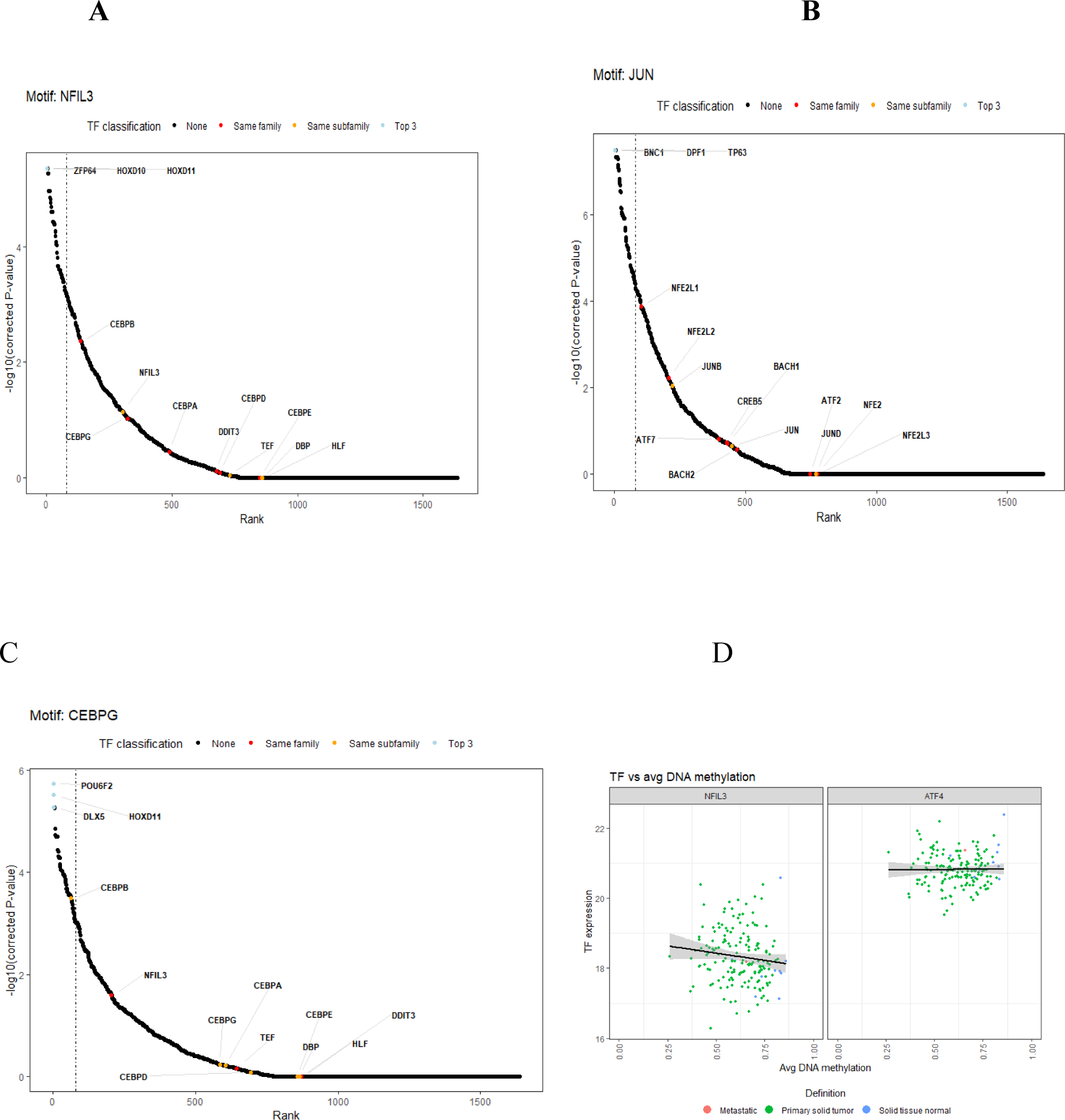
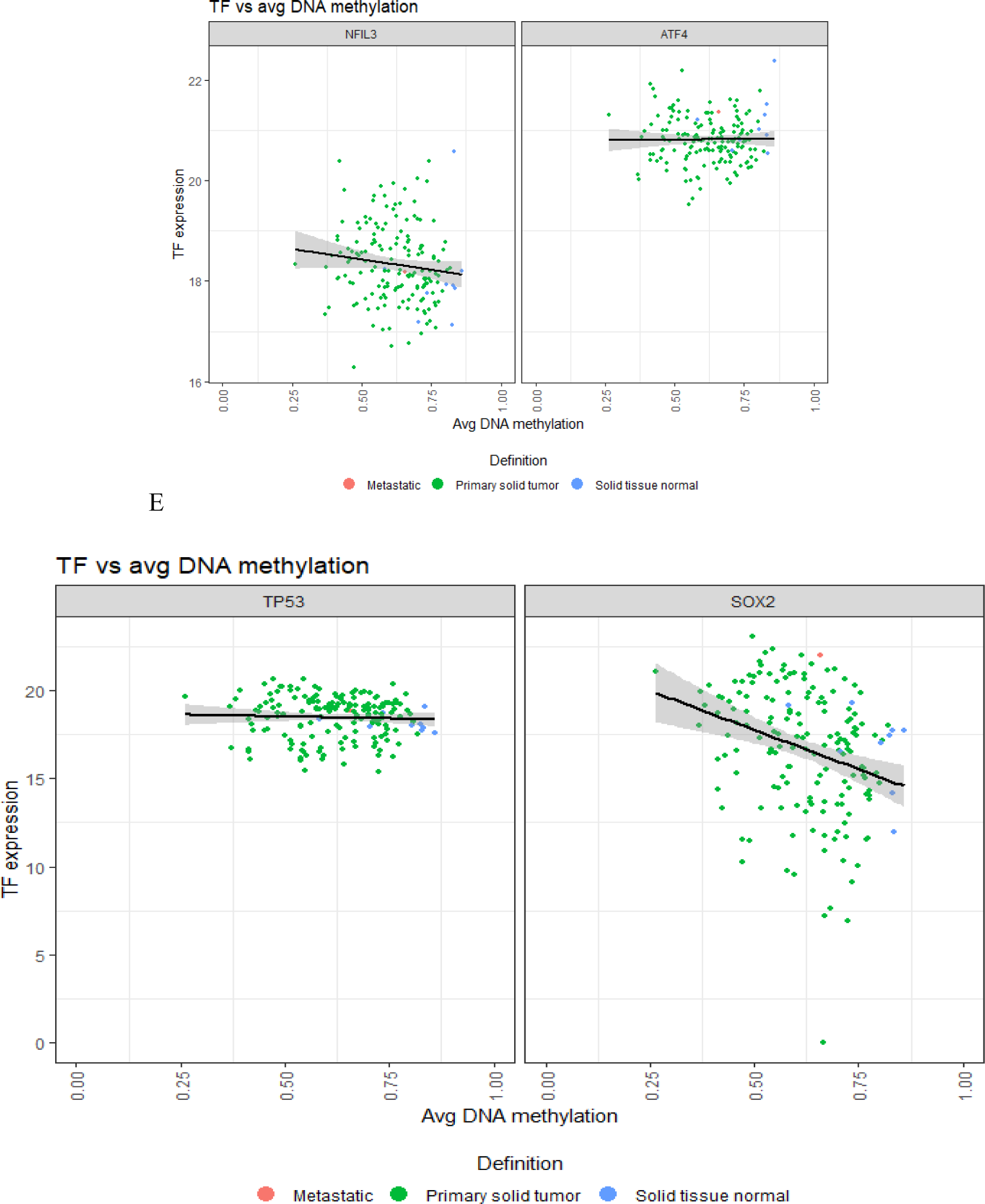
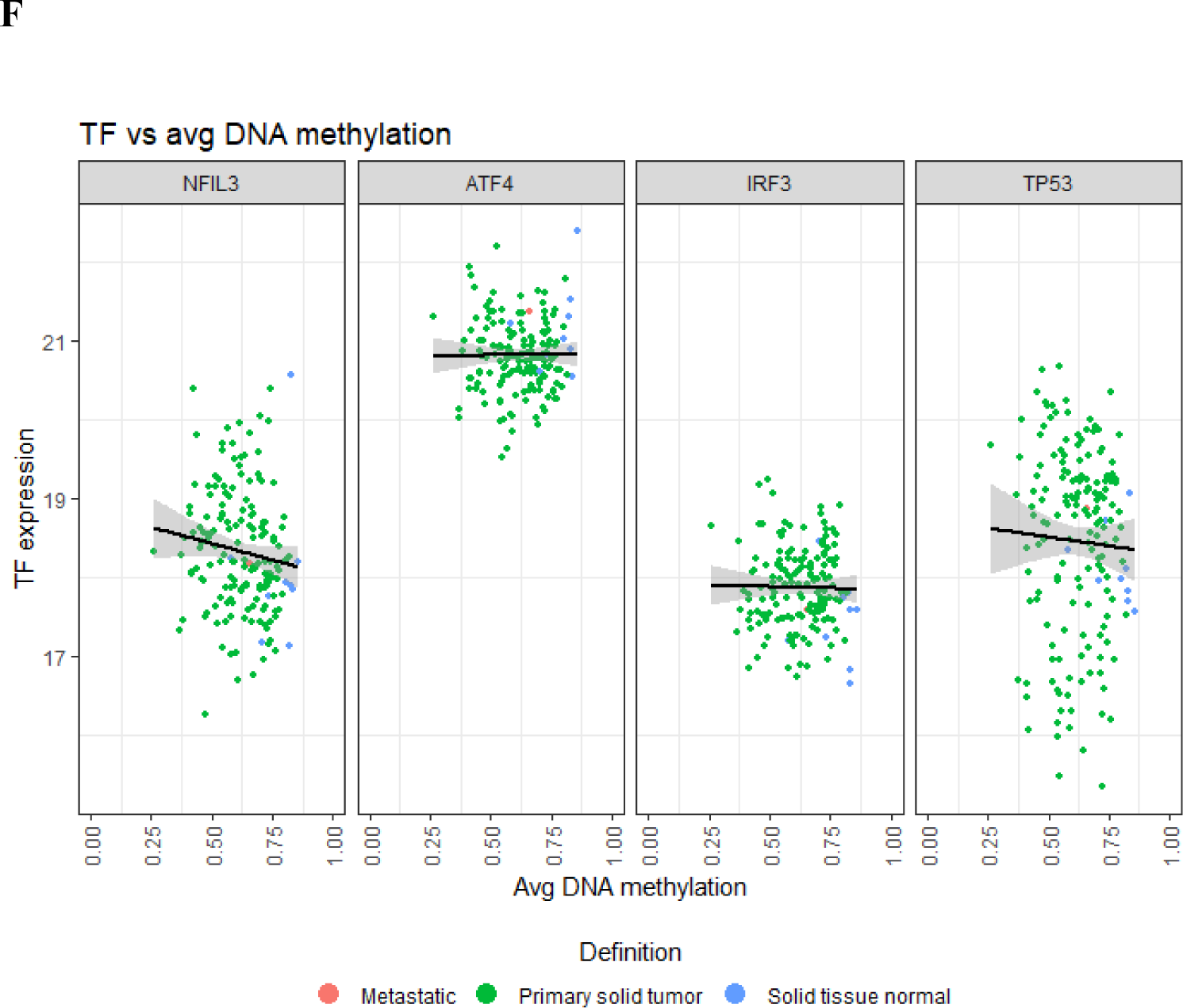
Transcription factors are classified through a comparative DNA methylation and TF expression analysis.

The NFIL3, JUN, and CEBPG motif exhibits a robust negative correlation, with NFIL3 being a promising candidate. Graphical representations in Figures A, B, and C emphasize transcription factors (TFs) in the same family (blue), subfamily (red), and NFIL3, JUN, and CEBPG (orange), respectively. Figure D illustrates the DNA methylation pattern of NFIL3 and ATF4 around TF expression. Plot E features the x-axis “TP53” and y-axis “SOX2”, with three-point types: “Metastatic tumor” (green circles with black outline), “Primary solid tumor” (solid green circles), and “Solid tissue normal” (green circles with white outline). The gray lines depict the best-fit trend among points. Scatter plot (F) compares TF expression and average DNA methylation across tissue types, with separate panels for Nfil3 and ATF4. The x-axis displays average DNA methylation, whereas the y-axis shows TF expression. The points are color-coded: green for metastatic tissue, blue for primary solid tumor, and gray for solid tissue normal. Each panel features a black trend line, demonstrating the effect of DNA methylation on gene expression across tissues.

**Table 4.4.**
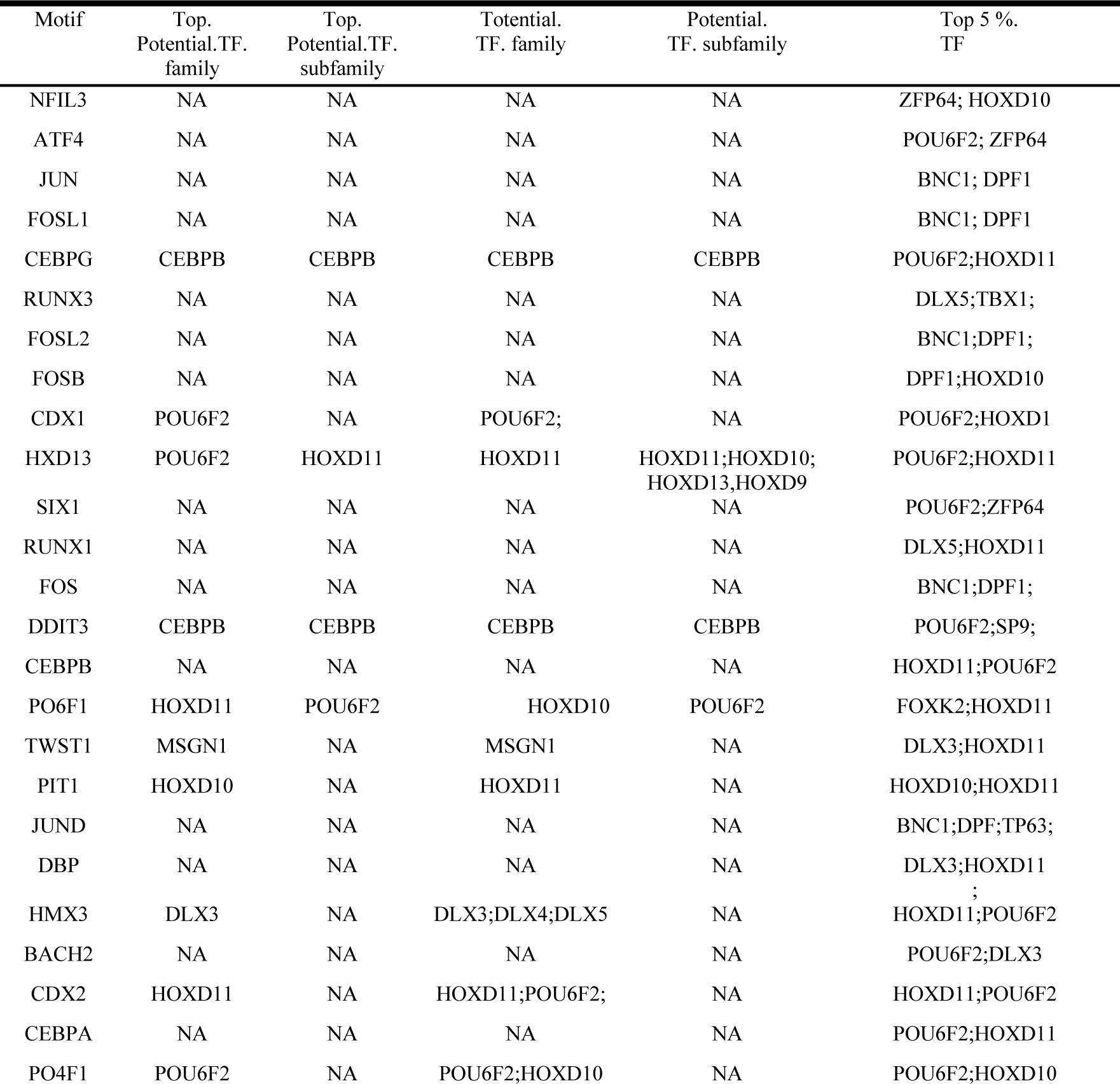
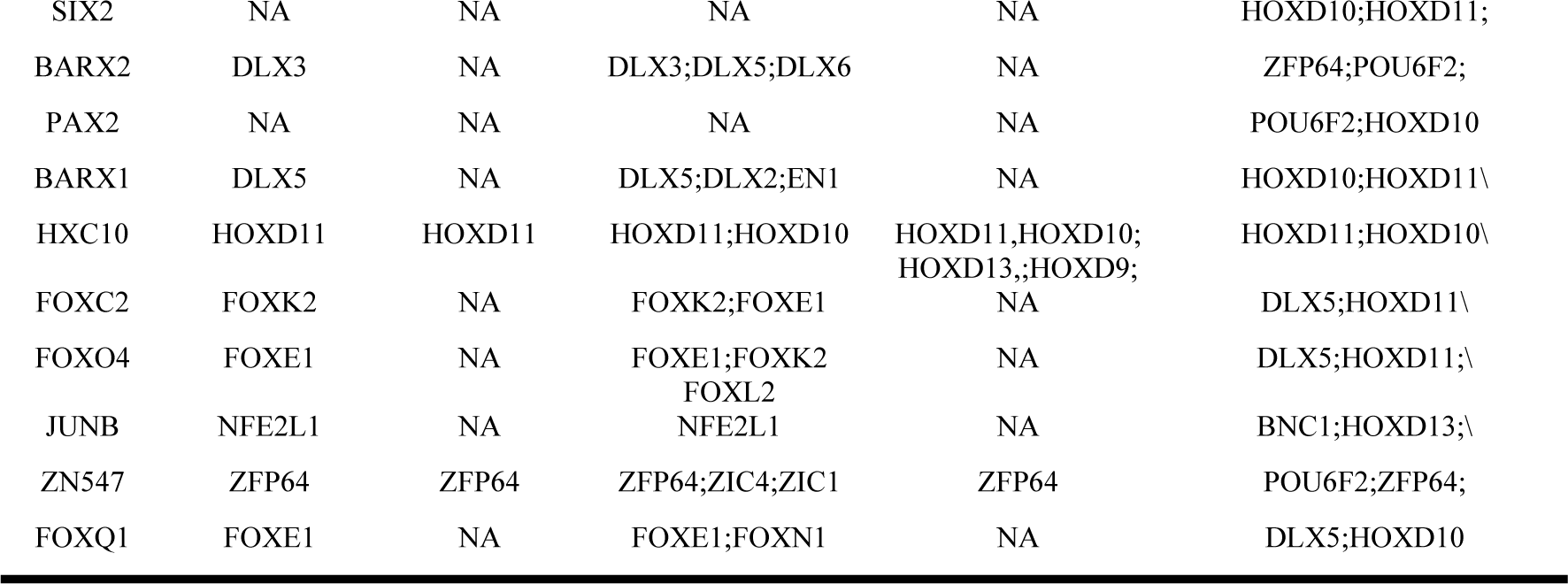
indicates the top 5 percent of the transcription family, potential transcription family, and top potential TF family, along with essential motifs obtained during analysis.

Table 4.5 displays the top 20 Human motifs from ESCA analysis, each linking to potential TF families and the top 5% of TFs. This aids researchers in more accessible ESCA analysis. The table lists enriched motifs, their corresponding TFs, family, and the highest-ranked TF within the family. TF subfamilies and top potential subfamilies align with TF subfamily classification. The analysis resulted in 35 TF motifs. The presented table discloses perceptive correlations between motifs and prospective TF families and subfamilies. To illustrate, motifs such as CEBPG_HUMAN H11MO.0.B and DDIT3_HUMAN H11MO.0.D are linked to the CEBPB TF family. Furthermore, the motif HXD13_HUMAN H11MO.0.D is affiliated with the HoxD11 and HoxD10 TF subfamilies. These associations suggest potential regulatory mechanisms that involve the motifs and the corresponding TF families and subfamilies. These findings are instrumental in untangling the intricate gene regulation networks and possible transcriptional control.

## 5 Survival analysis across multiple variables of ESCA

**Figure 5.1.**
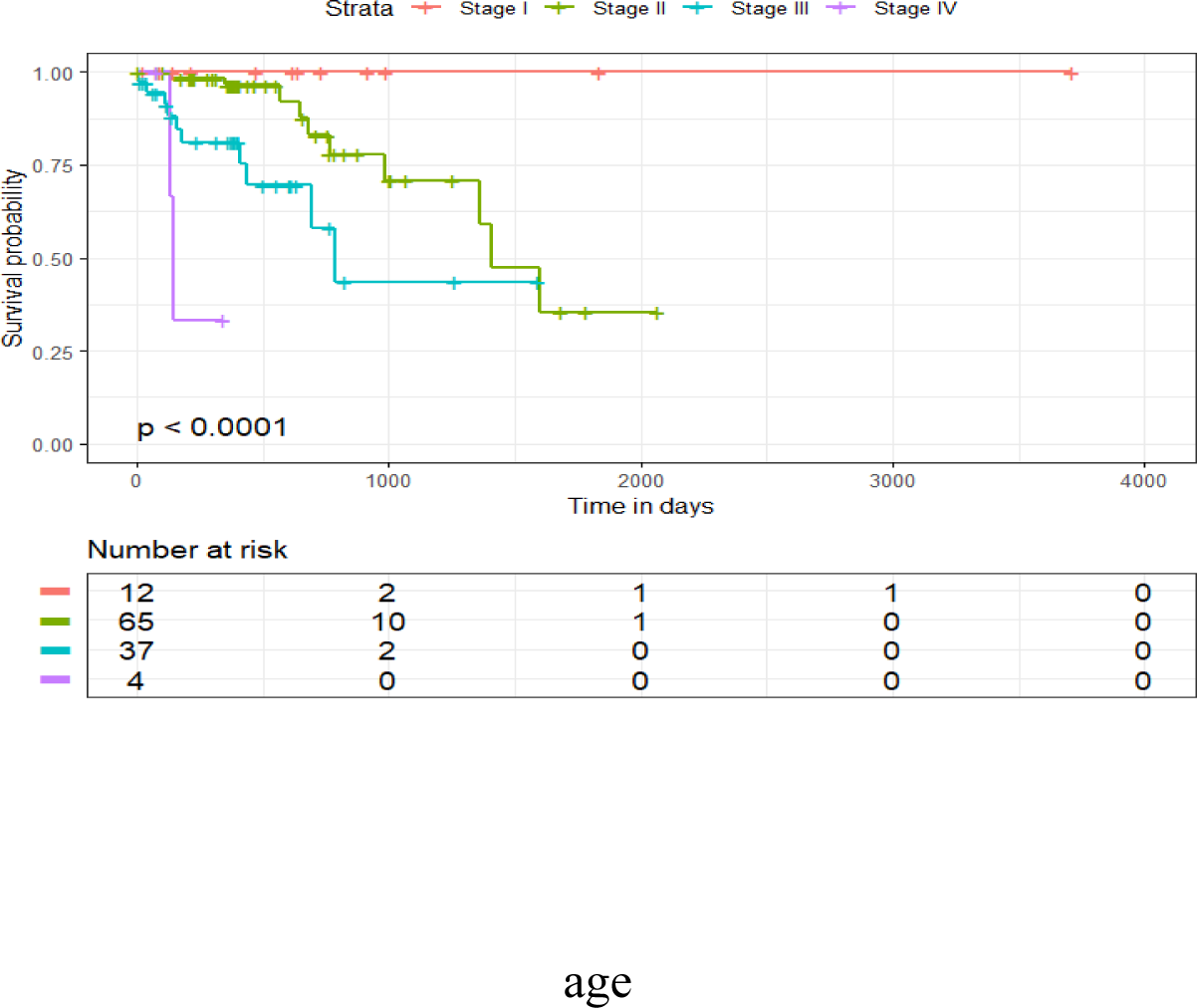
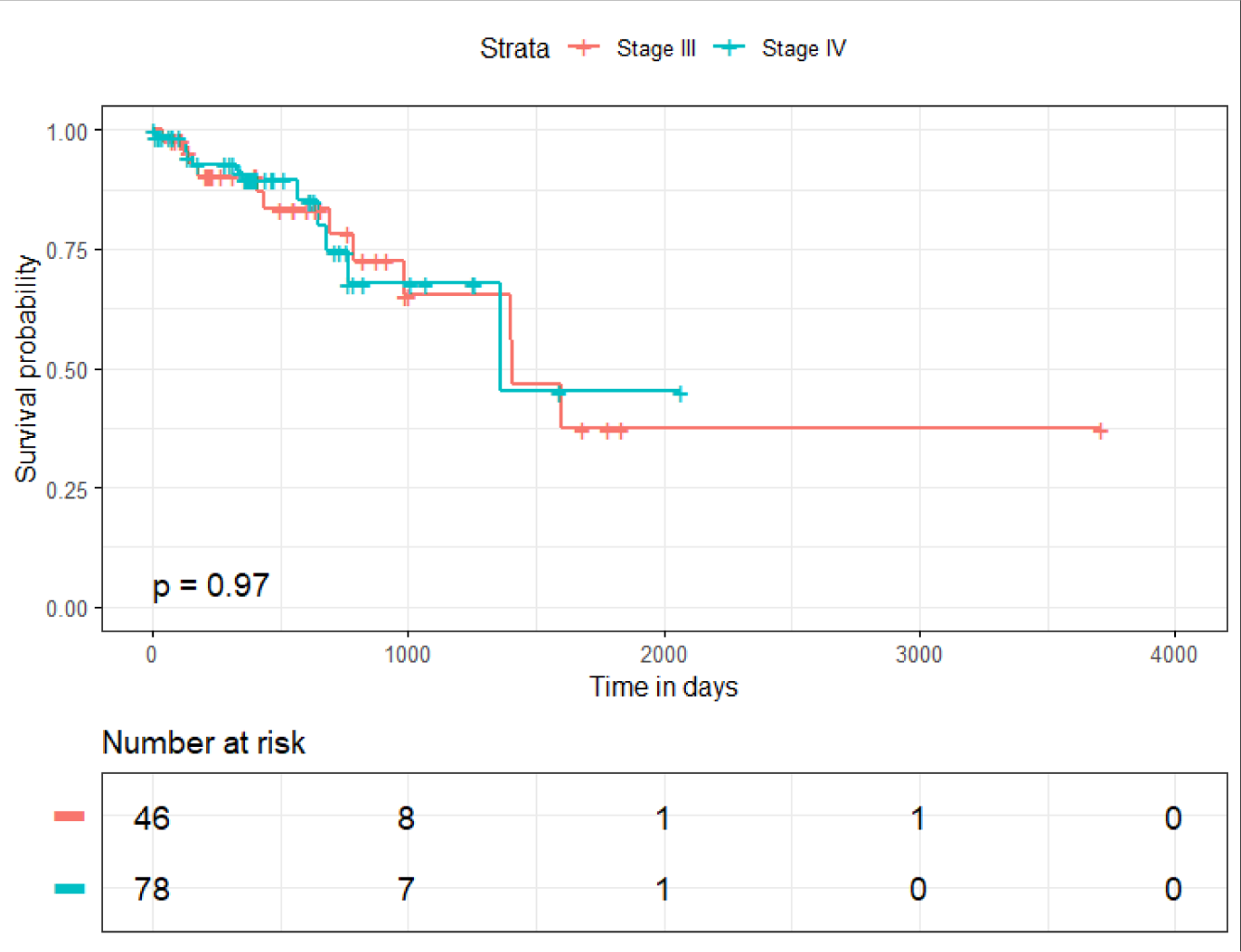
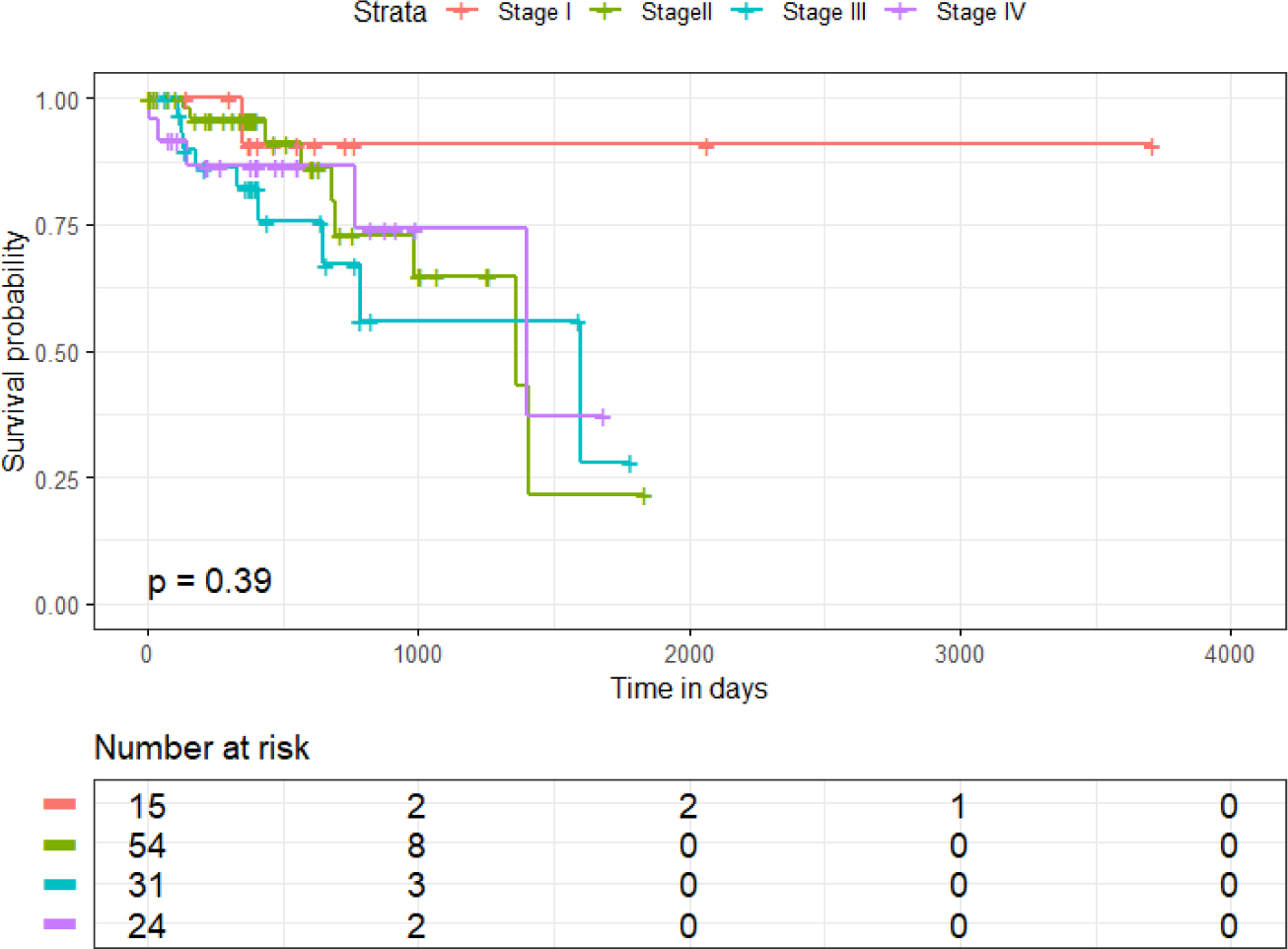
The graph illustrates the evolving survival probabilities at different cancer stages.

Fig: 5. 1 provides a visual illustration using Kaplan-Meier survival curves to illustrate the survival probabilities based on tumor stage, histological type, and pathological anatomy. Color-coded lines distinctly mark the different stages of diminishing survival probability, while an accompanying table enumerates the decreasing at-risk population. A striking difference in survival rates between stages becomes evident and is deemed statistically significant when the p-value is 0.0001 or lower. A highly significant p-value of less than 0.0001 is observed for the pathological tumor stage curve. In contrast, the survival probabilities for the histological type and patients’ pathological anatomy are p = 0.97 and p = 0.39, respectively, indicating non-significant differences.

## 6 Multivariate Analysis of ESCA Clinical Variables

The Cox proportional hazard model is utilized to conduct ESCA clinical variable analysis and determine the predicted survival function. The following section outlines the Cox regression of death time against the constant time variable, as demonstrated in Model 1 below.

### Model 1

**Table 6.1.**
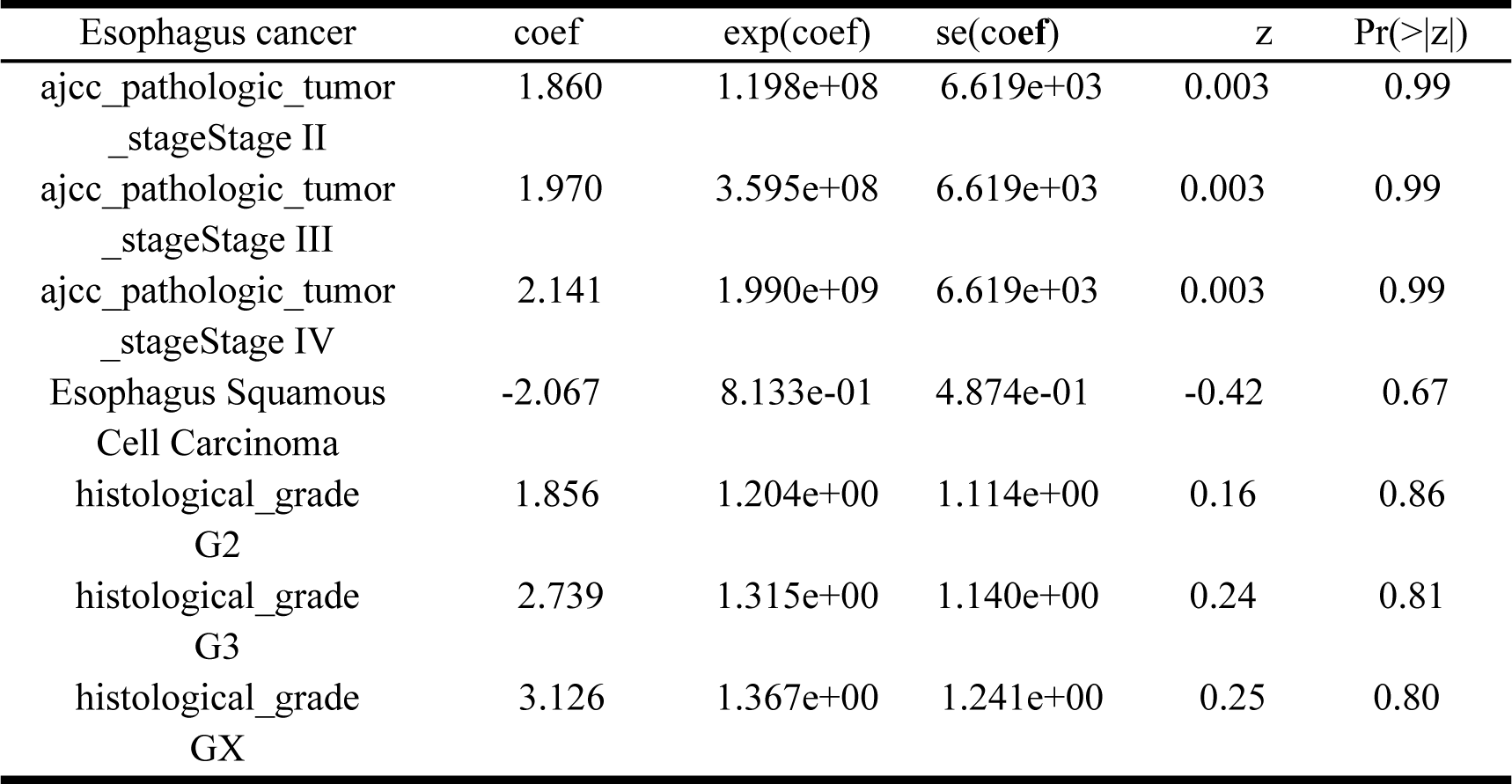
A tabular presentation of coefficients and statistical measures about variables linked to the analysis of esophageal cancer.

| Esophagus cancer | coef | exp(coef) | se(coef) | z | Pr(>\|z\|) |
| --- | --- | --- | --- | --- | --- |
| ajcc_pathologic_tumor<br>_stageStage II | 1.860 | 1.198e+08 | 6.619e+03 | 0.003 | 0.99 |
| ajcc_pathologic_tumor<br>_stageStage III | 1.970 | 3.595e+08 | 6.619e+03 | 0.003 | 0.99 |
| ajcc_pathologic_tumor<br>_stageStage IV | 2.141 | 1.990e+09 | 6.619e+03 | 0.003 | 0.99 |
| Esophagus Squamous<br>Cell Carcinoma | -2.067 | 8.133e-01 | 4.874e-01 | -0.42 | 0.67 |
| histological_grade<br>G2 | 1.856 | 1.204e+00 | 1.114e+00 | 0.16 | 0.86 |
| histological_grade<br>G3 | 2.739 | 1.315e+00 | 1.140e+00 | 0.24 | 0.81 |
| histological_grade<br>GX | 3.126 | 1.367e+00 | 1.241e+00 | 0.25 | 0.80 |

**Table 6.2.**
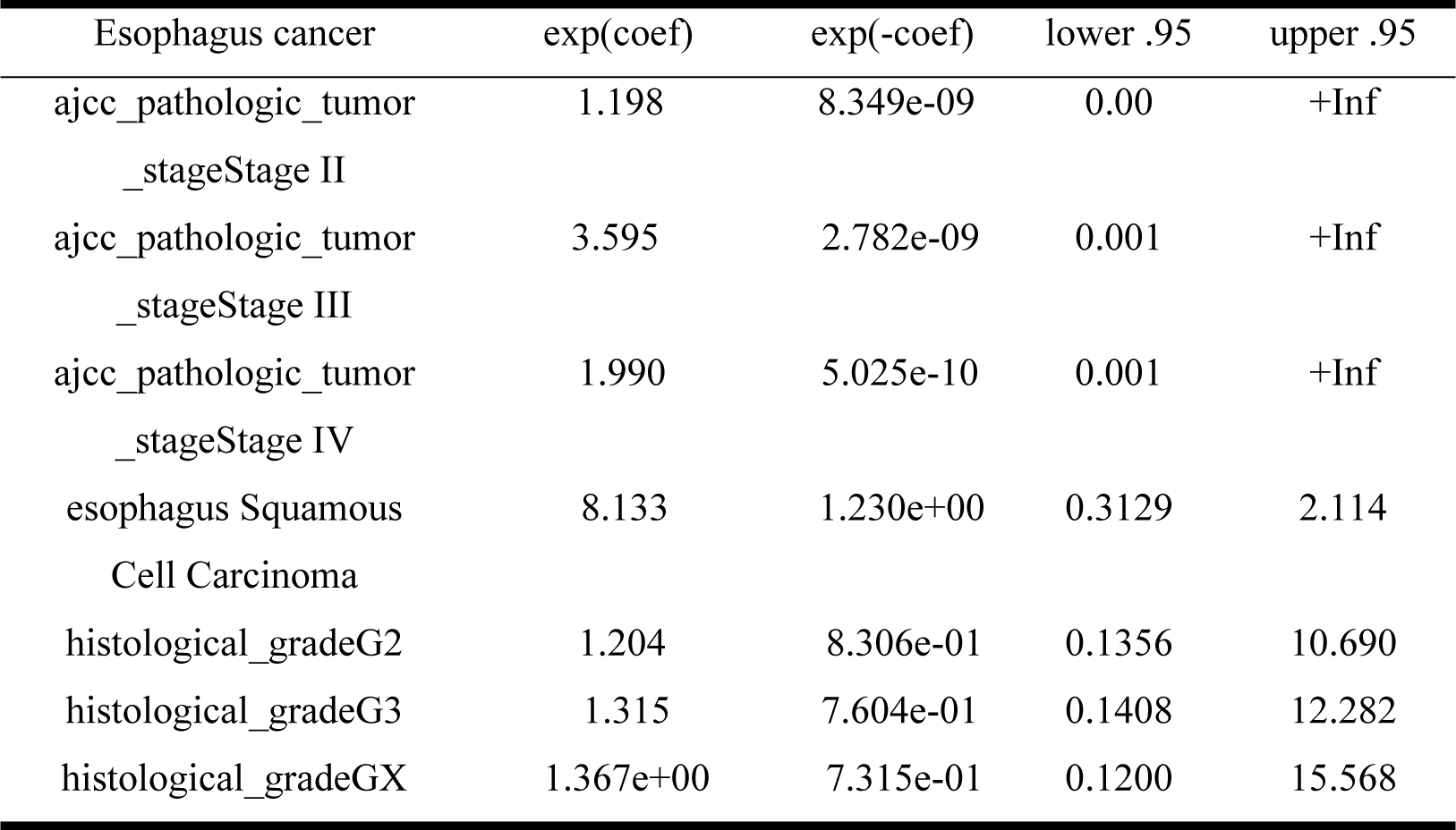
tabular presentation of coefficients and statistical measures about variables linked to the analysis of esophageal cancer.

A presentation of coefficients and statistics for variables related to esophageal cancer is depicted in a table. Noteworthy significance is observed in Stages II, III, and IV, where hazard ratios increase. While the risk for esophageal squamous cell carcinoma decreases, G2, G3, and GX grades exhibit minimal impact. The significance is indicated by p-values, and exponentiated coefficients are used to determine risks. Stages II, III, and IV demonstrate advantageous effects with exponentiated coefficients ranging from 1^8 to 1^9. Specific categories of Histological Grade show minor positive effects. The Cox proportional model for ESCA clinical variables is presented in the table, where positive coefficients suggest an amplified mortality probability, except for Esophagus Squamous Cell Carcinoma, indicating a reduced risk. The risk increases from Stage II to IV, with exponential coefficients indicating risk ratios. The confidence intervals range from 0.3129 to 2.114. P-values are crucial for probability, Wald, and score tests, and in model 1, p-values of 0.009, 0.07, and 8e-04 signify its significant impact.

**Figure 6.1.**
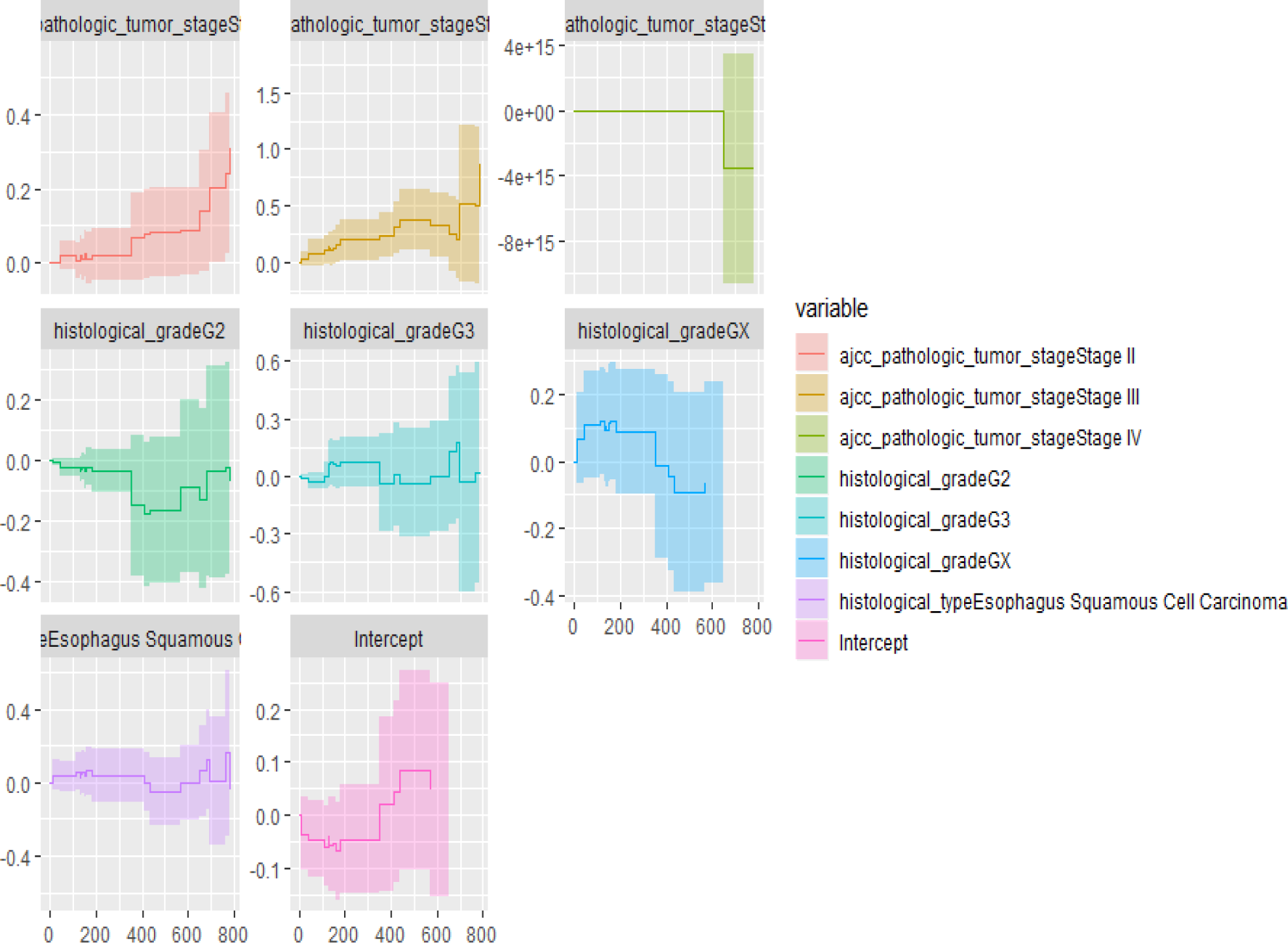
Alen regression additive model.

The Aareg function, Alen regression additive model, is used to analyze the correlation between histologic tumor stages and histologic grade in Esophageal Squamous Cell Carcinoma. Examining the graph helps medical professionals and researchers make better treatment decisions and predict patient outcomes.

### 6.1 Assessing proportional Hazards assumption

The study elaborates on the Cox risk model’s techniques for validating hypotheses. Three diagnostic forms for the Cox model are analyzed: assessing the proportional hazard hypothesis, identifying adequate perception, and detecting nonlinearity. The assumption is evaluated via 30 and visual checks with Schoenfeld residuals. Schoenfeld residue measurements and ggcoxzph can Table 6.3: The values presented herein reveal the results of utilizing a chi-square test for association with the various ESCA clinical variables. Upon conducting the said test, it has been detected that there exists a marked and remarkable association between the histological_grade and GLOBAL variables, as evidenced by the p-values that are initiated to be less than 0.05. Conversely, no statistically significant association between the ajcc_pathologic_tumor_stage and histological_type variables can be established.

**Table 6.3.** Chi-square test for association about the various ESCA clinical variables.

| ESCA clinical variables | Chisq | df | p |
| --- | --- | --- | --- |
| ajcc_pathologic_tumor_stage | 4.797 | 3 | 0.187 |
| histological_type | 0.612 | 1 | 0.434 |
| histological_grade | 11.331 | 3 | 0.010 |
| GLOBAL | 15.887 | 7 | 0.026 |

**Figure 6.2.**
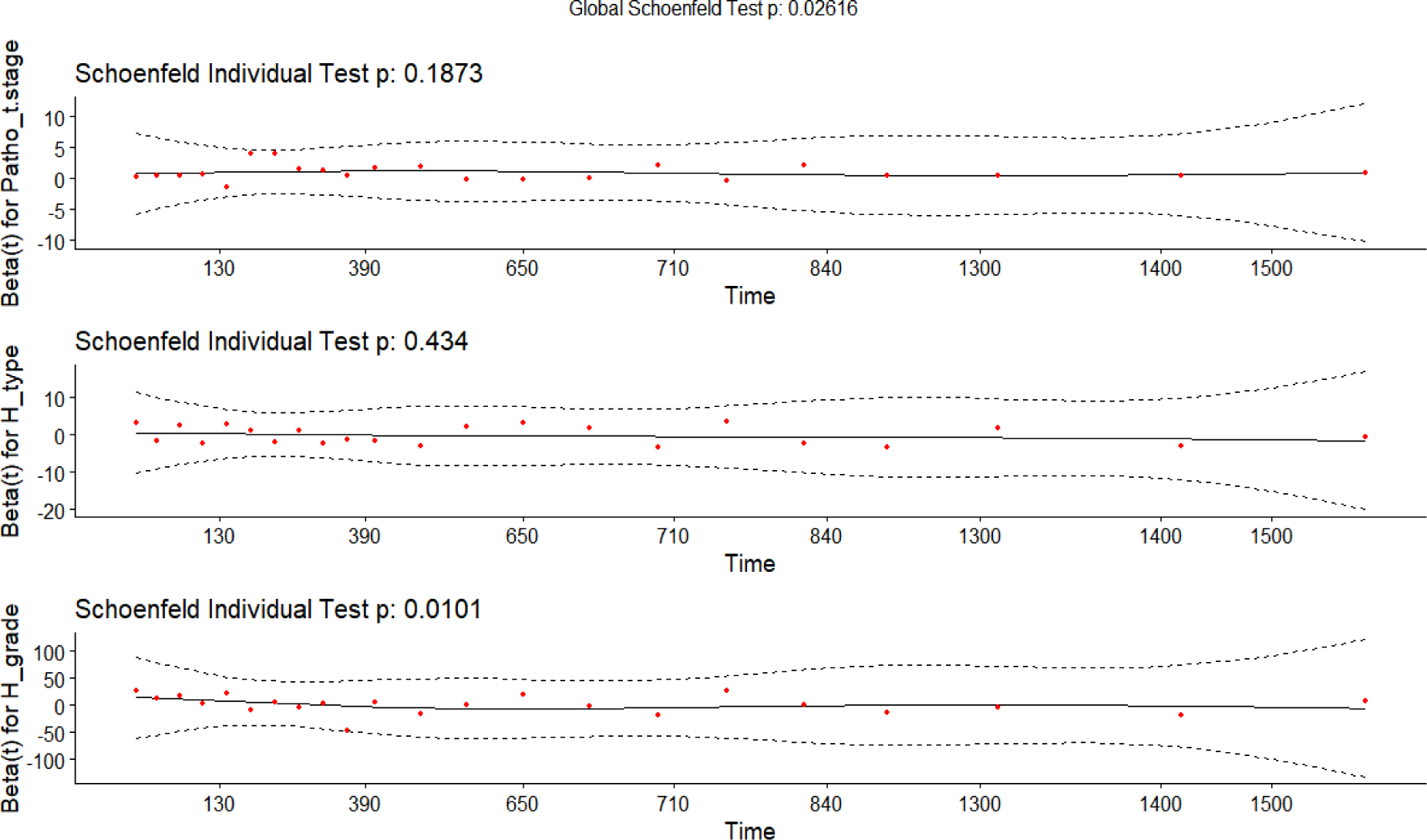
Schoenfeld Individual Test.

The graphs presented in the study have been furnished with descriptive titles, which are as follows: “Schoenfeld Individual Test: p = 0.1873,” “Individual Test: p = 0.434,” and “Schoenfeld Individual Test: p = 0.0101.” These diagrams symbolize the results derived from diverse iterations of a Schoenfeld individual test, each bearing unique and linked p-values. The compilation of illustrations effectively showcases Schoenfeld’s evaluations, with each evaluation corresponding to a distinct p-value. The horizontal axis is representative of duration, while the vertical axis is aligned with the beta coefficient for the logarithm of HR and its standard error. Red indicators strategically placed along the dotted lines have facilitated the identification of significant points.

### 6.2 Testing influential observations

**Figure 6.3.**
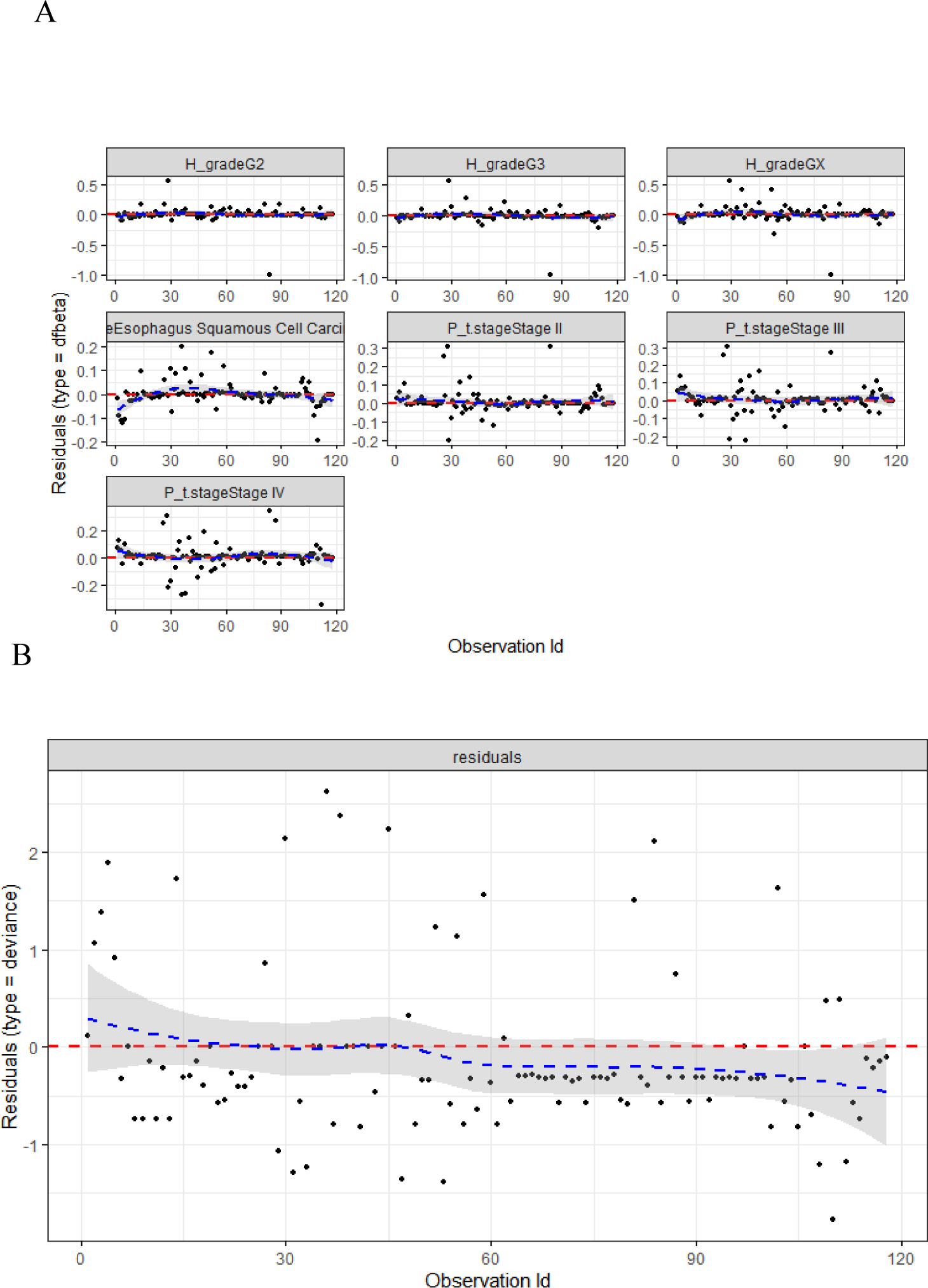
Cox Regression beta Plot for Modeling Time to Death.

Figure 6.3 shows a plot for Cox Regression beta for time-to-death modeling. The index diagram displays dfbeta values for Cox regression over time. Deviance residuals are highlighted in Figure 6.3 B to show differences between observations and deviance. Index 6.3: A showcases beta values for Cox’s time-to-death regression involving Pathological_tumor_stage, histological_type, and histological_grade. Figure 6.3 A displays residuals on the Y-axis and linear estimates or observation indices on the X-axis. The high-index (6.3: A) graph reveals incongruities in specific dfbeta values that impact regression coefficients. Diagram 6.3: B displays the likelihood of detecting abnormal values via deviation residues, suggesting a hypothesis for survival predictions.

## 7 Leveraging Reverse Phase Protein Array (RPPA) Protein-Level Data for In-depth Analysis

**Table 7.1.**
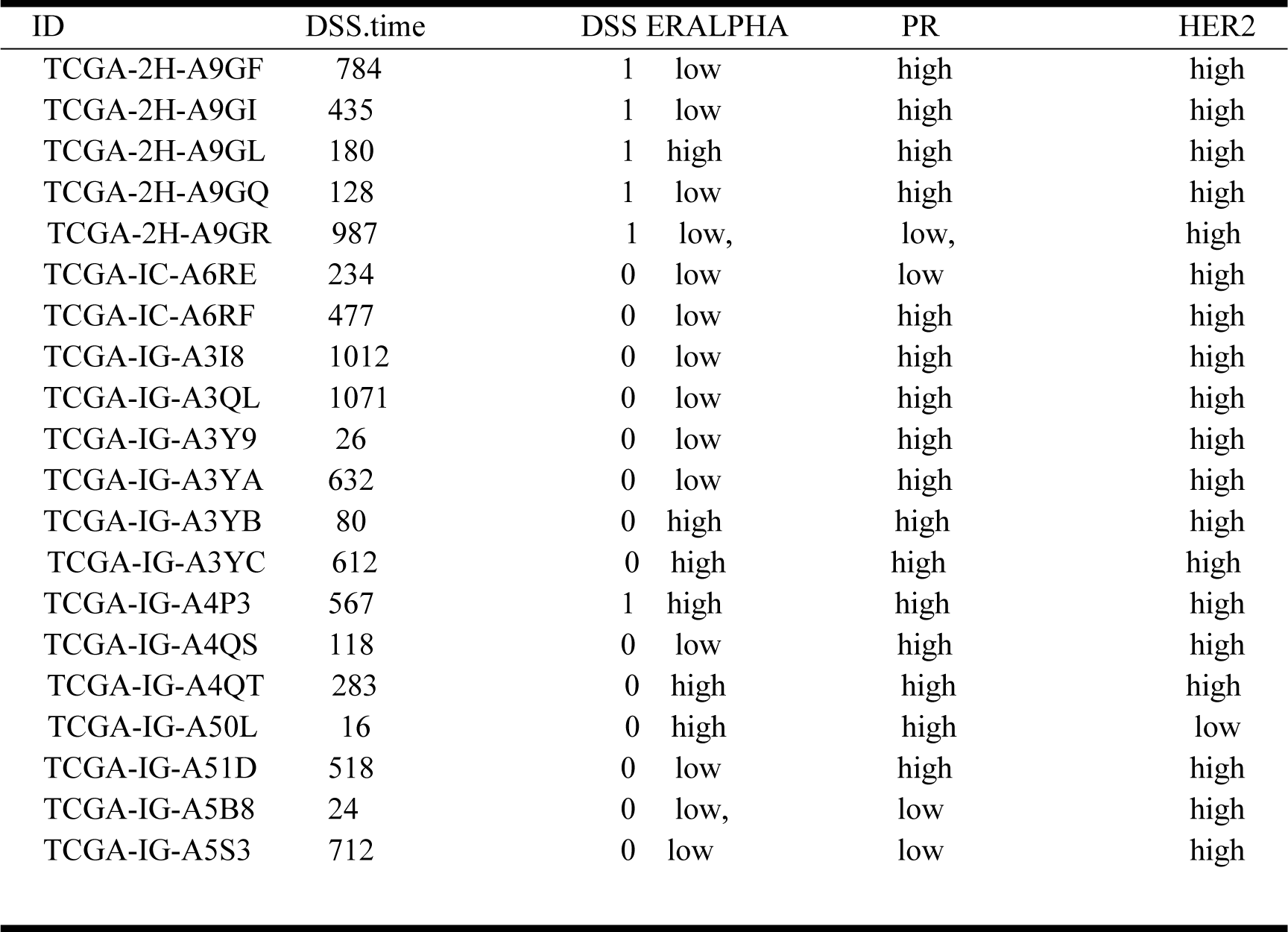
(RPPA) Protein-Level Data for In-depth Analysis.

Clinical variables are straightforward once derived from multivariate analysis; they are categorical. We must recognize the proteins of interest, identify the pieces, and convert them into categorical variables. We selected proteins tested before through immunohistochemistry, a part of the standard procedure for diagnosing breast cancer and treatment. However, we used these three types of proteins to analyze ESCA clinical variables. These proteins are ERALPHA, PR, and HER2.

Table 7.1 displays protein expression with ESCA clinical variable. A study examines various elements in distinct samples. Columns demonstrate time measurements and outcomes. Protein markers disclose marker levels. Each row corresponds to a unique sample. HER2 consistently manifests a high level across samples. Anticipating the implications of the HER2 marker could yield valuable insights.

**Table 7.2.** A Prognostic Model for Esophageal Cancer (ESCA).

|  | **coef** | **exp(coef)** | **se(coef)** | **z** | **Pr(>\|z\|)** |
| --- | --- | --- | --- | --- | --- |
| **ERALPHA (low)** | 8.092e-01 | 4.452e-01 | 4.962e-01 | 1.631 | 0.103 |
| **PR (low)** | 5.825e-01 | 5.585e-01 | 6.697e-01 | 0.870 | 0.384 |
| **HER2 (low)** | 1.412e+00 | 2.436e-01 | 1.046e+00 | 1.350 | 0.177 |
| **tumor_stageStage II** | 1.884e+01 | 1.522e+08 | 6.638e+03 | 0.003 | 0.998 |
| **tumor_stageStage III** | 1.994e+01 | 4.572e+08 | 6.638e+03 | 0.003 | 0.998 |
| **tumor_stageStage IV** | 2.124e+01 | 1.681e+09 | 6.638e+03 | 0.003 | 0.997 |
| **histological type Esophagus**<br>**Squamous Cell Carcinoma** | 3.391e-01 | 7.124e-01 | 4.431e-01 | 0.765 | 0.444 |

**Table 7.3.**
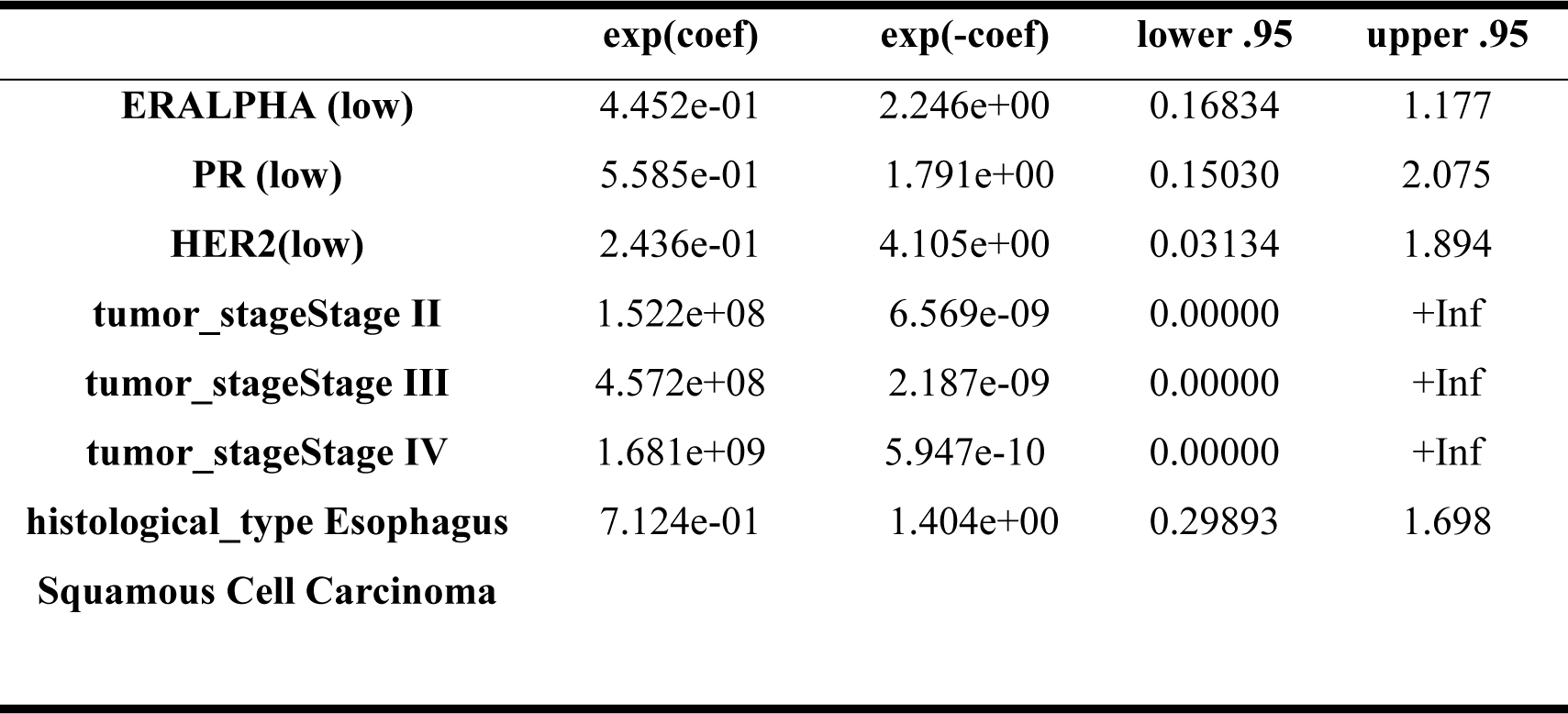
B Prognostic model for Esophageal Cancer (ESCA).

|  | **exp(coef)** | **exp(-coef)** | **lower .95** | **upper .95** |
| --- | --- | --- | --- | --- |
| **ERALPHA (low)** | 4.452e-01 | 2.246e+00 | 0.16834 | 1.177 |
| **PR (low)** | 5.585e-01 | 1.791e+00 | 0.15030 | 2.075 |
| **HER2(low)** | 2.436e-01 | 4.105e+00 | 0.03134 | 1.894 |
| **tumor_stageStage II** | 1.522e+08 | 6.569e-09 | 0.00000 | +Inf |
| **tumor_stageStage III** | 4.572e+08 | 2.187e-09 | 0.00000 | +Inf |
| **tumor_stageStage IV** | 1.681e+09 | 5.947e-10 | 0.00000 | +Inf |
| **histological_type Esophagus**<br>**Squamous Cell Carcinoma** | 7.124e-01 | 1.404e+00 | 0.29893 | 1.698 |

The tables provide profound insights into a prognostic Esophageal Cancer (ESCA) model. Specifically, Table 7.3A proffers coefficients and significance levels for pertinent factors, such as protein markers (i.e., ERALPHA, PR, and HER2), tumor stage categories (II, III, and IV), and histological type (Esophagus Squamous Cell Carcinoma). Tumor stage categories carry substantial coefficients, suggesting their potential impact. Table 7.3: B also provides exponentiated coefficients and confidence intervals, which aid direct odds ratio interpretation. The significant outcomes observed in both tables emphasize strong associations with ESCA prognosis, particularly those pertaining to tumor stage categories.

**Table 7.4.** Statistical test.

|  |  |  |  |
| --- | --- | --- | --- |
| Concordance |  | = 0.8 (se = 0.051) |  |
| Likelihood ratio test | = 25.41 | on 7 df, | p=6e-04 |
| Wald test | = 17.77 | on 7 df, | p=0.01 |
| Score (log rank) test | = 30.74 | on 7 df, | p=7e-05 |

Table 7.4 presents noteworthy discoveries: the moderate agreement between anticipated and observed outcomes (0.8), the superiority of the intricate model demonstrated by the Likelihood Ratio Test (25.41 statistics, p < 0.001), the importance of the coefficient affirmed by the Wald Test (17.77 statistics, p = 0.01), and apparent differences in survival distribution uncovered by the Score Test (30.74 statistics, p < 0.0001). These outcomes emphasize significant relationships and contradictions within the study.

## 4. Discussion

Esophageal cancer (ESCA) remains a significant worldwide health issue, necessitating a profound comprehension of its genetic complexities to enhance diagnosis and treatment approaches. 31 Consequently, Early diagnosed patients with ESCA cancer will have a longer life expectancy and lower mortality. This research sheds light on the genetic mechanisms governing ESCA and highlights the potential clinical impact of early diagnosis [34]. The study’s central aim was to decipher ESCA’s genetic underpinnings by meticulously analyzing gene expression quantification, RNA expression, DNA methylation, mutation, and clinical data from the TCGA database through the Bioconductor repository[35]. The researchers utilized an assortment of analytical methodologies to reveal valuable perceptions, comprising the recognition of expressed genes through the utilization of the edgeR application, controlling multiple testing using the Benjamini & Hochberg methodology[36], and employing diverse R-based visualization tools, such as cluster plots and heat maps, for data representation [29]. These tools collectively enhanced the understanding of intricate genetic patterns that underpin ESCA. The study’s influence of changed DNA methylation on gene expression is a crucial finding. Our research team uncovered that changed DNA methylation in tumors can influence gene behavior, silencing suppressor genes while activating oncogenes through hyper/hypomethylation. This shift in gene expression dynamics potentially plays a crucial role in ESCA progression. The study utilized the unsupervised machine-learning Bioconductor packages, leveraging unsupervised machine-learning techniques to unravel DNA methylation and RNA expression patterns) By employing this approach, the researchers successfully identified hypo/hypermethylated regions of interest associated with ESCA, discriminating between tumor and regular groups and highlighting significant probe-gene pairs. This methodological innovation enhances our understanding of ESCA’s intricate genetic makeup and its implications for patient outcomes.

A notable contribution of this study lies in its innovative use of statistical methodologies. Through a statistical 20-n test approach, the researchers dissected the regulatory mechanisms behind enhancer probes exhibiting differential methylation patterns in ESCA. By investigating downstream and upstream genes linked with each probe-gene pair, they could pinpoint specific hypomethylated values, shedding light on the regulatory pathways driving ESCA’s genetic aberrations. Moreover, Fisher’s confidence test was employed to identify enriched motifs associated with these regulatory mechanisms, yielding significant insights into transcription factor interactions. The study’s clinical relevance is underscored by its prognostic analyses. Utilizing the Cox proportional hazard statistical model, the researchers were able to make predictive forecasts for clinical variables in ESCA. The Schoenfeld analysis test and Reverse Phase Protein Array (RPPA) analysis provided additional layers of insight into the clinical implications of the genetic findings [37]. This holistic approach not only expands our understanding of the genetic basis of ESCA but also has the potential to guide personalized treatment decisions and prognosis.

In summary, this investigation thoroughly investigates the hereditary terrain of esophageal cancer, highlighting the significance of DNA methylation modifications in molding the dynamics of gene expression. By utilizing an assortment of innovative analytical methods and statistical approaches, the team of researchers exposes pivotal genes, pathways, and regulatory mechanisms that contribute to the advancement and progression of ESCA. These discoveries enhance our comprehension of illness and offer the potential for creating targeted therapies and improved clinical outcomes for patients diagnosed with ESCA.

## Conclusion

This research elucidates the crucial association between DNA methylation and esophageal cancer (ESCA), highlighting its significance as a pivotal biomarker for predictive and diagnostic purposes. Through implementing an unsupervised machine learning algorithm based in R, the study reveals distinct DNA methylation patterns in ESCA tumors compared to normal tissue, providing a promising avenue for effective cancer diagnosis. The investigation’s identification of enriched transcription factor binding motifs associated with target genes illuminates gene expression mechanisms and classification within ESCA, thereby offering deeper insights into its underlying dynamics. This study underscores the potential of DNA methylation analysis, in conjunction with advanced artificial intelligence techniques and methylome-based approaches, to enhance early detection and prediction of ESCA, consequently leading to novel biomarkers and improved patient outcomes for esophageal cancer and beyond.

## Data Availability

Upon the acceptance of this manuscript for publication, the supporting data and associated code will be made publicly available on GitHub for the benefit of the research community. Interested researchers can access these resources at the following repository: https://github.com/aliswatian1989.

https://www.cancer.gov/

## CONFLICTS OF INTEREST

The authors declare that they have no conflicts of interest.

### Ethical approval

This article does not contain any studies with humans. Participants or animals performed by any of the authors.

**Informed consent** was obtained from all individual participants included in the study.

## Acknowledgments

Self-funded research work.

